# Common and distinct neurofunctional signatures of emotion regulation strategies and their clinical translation in dynamic naturalistic contexts

**DOI:** 10.1101/2025.05.29.25328539

**Authors:** Heng Jiang, Jingxian He, Kaeli Zimmermann, Xinqi Zhou, Xianyang Gan, Stefania Ferraro, Lan Wang, Bo Zhou, Liyuan Li, Keith M Kendrick, Weihua Zhao, Dezhong Yao, Tifei Yuan, Feng Zhou, Benjamin Becker

## Abstract

Adaptive emotion regulation (ER) is essential for mental health. Reappraisal and acceptance are effective ER strategies that form the basis of therapeutic interventions, yet their common and distinct neural signatures - and the translational potential of these signatures - remain unclear. Here, we combined naturalistic fMRI with multivariate predictive modeling to develop neurofunctional signatures that accurately and comprehensively characterize negative affect and its regulation via acceptance and reappraisal in dynamic, immersive contexts (n=59). These signatures demonstrated process-specificity and generalization across cohorts, cultures, and modalities (n=33, n=358, n=33). ER strategies were encoded in distributed, distinguishable neural representations, with shared contributions from the default mode network and strategy-specific contributions from the amygdala, somatomotor and attention (acceptance), and the frontoparietal control (reappraisal) networks. The neuromarkers precisely identified strategy-specific ER impairments in cannabis users (n_healthy_controls_=48, n_cannabis_users_=49), underscoring their translational relevance. We provide comprehensive, clinically-relevant brain models of emotion regulation and dysregulation in naturalistic contexts.

## Introduction

Emotion regulation (ER) – the ability to modulate the experience and expression of emotions – is fundamental to adaptive functioning and mental health^1–6^. Impairments in ER represent a core transdiagnostic feature across major mental disorders (MDs)^5,7–9^. Regulation of negative affect can be achieved by several mental strategies, with cognitive reappraisal and acceptance being particularly effective^2,10–13^. Both are central building blocks of psychotherapeutic interventions^14–16^ and have a high potential of being applied in everyday emotional challenges^17,18^.

While both strategies efficiently regulate negative affect and focus on cognitive changes^13^ (see *process model of emotion regulation*^1,2^), they differ markedly in their operational and cognitive mechanisms. Reappraisal is based on reinterpreting the subjective meaning of the emotion-inducing event to control its affective impact^19,20^. Its successful implementation critically relies on an array of executive functions, including attention, working memory, and cognitive control, and represents an active and cognitively demanding process^21,22^. In contrast, acceptance involves a nonjudgmental awareness of emotional experiences without attempts to control them^23,24^. As such, it is considered a less cognitively demanding and more passive strategy relying on self-referential processes or metacognitive awareness^22^.

Neurobiological models distinguish between brain systems involved in emotional reactivity – such as the amygdala and insula, and systems involved in regulation, including lateral prefrontal and parietal regions^25–27^. In line with the effectiveness of both strategies, fMRI meta-analyses indicate that both effectively modulate activity in subcortical emotion “reactivity” regions, such as the amygdala,^24,28^ while enhancing the engagement of ventrolateral prefrontal regulatory regions (vlPFC)^29,30^. Notably, reappraisal has been strongly associated with engagement of the fronto-parietal control network^26,27,31^, reflecting its reliance on regulatory cognitive processes, while acceptance has been associated with an engagement of core regions of the posterior midline default mode network (DMN)^22,24,32–34^, suggesting a distinct neural signature reflective of self-related and introceptive processing.

Despite extensive work using conventional neuroimaging and meta-analytic approaches several key questions remain unresolved, including (1) the common and distinct neural representations of reappraisal and acceptance remain controversially debated^22,35–37^, and, (2) the generalizability of the identified neural systems that have been extensively assessed using sparsely presented static stimuli (e.g. pictures) to naturalistic, dynamic emotional experiences which more closely reflect real-world affective experiences, as well as (3) the translational potential of these neural representations as precise neuromarkers for ER impairments in clinical populations.

First, traditional mass-univariate analysis, widely used in previous studies, presents several limitations with respect to establishing comprehensive and accurate brain models for specific mental processes, including (1) voxel-wise independence assumptions that reduce sensitivity to comprehensively determine cognitive functions^38^ and (2) averaging across voxels – each containing a massive amount of neurons (∼5.5 million) – yields nonspecific signals^39,40^. Recent advances in multi-variate predictive models convergently suggest that emotional and cognitive processes are encoded in distributed neural representations rather than in isolated brain systems^28,41–43^.

Second, few studies^22,33^ have directly compared acceptance and reappraisal strategies using a within-subject design or under ecologically valid conditions. Most neurobiologically-informed models instead rely on meta-analyses^5,44,45^, which are limited by the univariate approach of the original studies and further constrained by variability indicated by preprocessing protocols^46^. Moreover, these studies used sparsely presented, static, and isolated stimuli (e.g., affective pictures), which limits ecological validity^47,48^, in particular in the context of emerging evidence indicating that neural systems that support affective experience differ substantially when engaged during dynamic naturalistic experiences^49–51^.

Third, despite the recognition of impaired ER as a transdiagnostic impairment in MDs, including addiction^7,52,53^, findings have yet to be translated into clinically useful neuromarkers. While initial progress has been made in developing a neuromarker for cue reactivity, parallel progress in the domain of ER is lacking^54^.

To address these critical limitations, we developed and extensively evaluated a comprehensive, ecologically valid, and clinically applicable brain model of ER under naturalistic conditions. We here capitalize on the combination of functional magnetic resonance imaging (fMRI) with machine-learning-based multivariate pattern analysis (MVPA) and naturalistic paradigms^55–58^. MVPA leverages information across multiple spatial scales^39,59^, allowing the establishment of comprehensive, accurate, and generalizable whole-brain models of emotional experiences (‘neuroaffective signatures’) with improved sensitivity and specificity and translating these into neuromarkers for MDs^59,60^. To further increase the ecological validity of these models, recent studies have employed naturalistic paradigms with dynamic videos, speech, and music as materials that are consistent with the typical sensory stimuli encountered in realistic daily life^47–49,61,62^.

Against this background, we aimed here at precisely determining common and distinct neural representations of reappraisal and acceptance during naturalistic dynamic emotional processing combined with MVPA-based neural decoding. Briefly, 59 participants from a pre-registered discovery cohort and 33 participants from a pre-registered validation cohort underwent a dynamic naturalistic ER paradigm with concomitant fMRI (Fig.1a), with acceptance (NA), reappraisal (NR), or natural emotional experience to negative (NV) or neutral clips (NeutV). We next systematically tested (1) whether it is possible to develop precise and robust neural signatures to determine negative experiences (NNES) and their regulation via reappraisal and acceptance (NERS-R or NERS-A, respectively) across the discovery cohort and independent validation cohort (Fig.1b); (2) whether the developed decoders generalize to independent ER processing investigated using conventional static paradigms (e.g., picture) in a large dataset of healthy individuals (n=358)^28^ and reappraisal pain induced by heat stimuli (n=33)^63^. Combing multiple analysis methods to determine (3) the distinct and common brain representations and brain systems that underly acceptance and reappraisal, utilizing e.g. Bayes Factor (BF) analysis which allows examining for both the null and alternative hypotheses ^28,64,65^ and spatial similarity analysis with encoding models and prefrontal systems involved in ER^66^ as well as large-scale functional networks^67^ (Fig.1c). Based on accumulating evidence for ER deficits in MDs, including excessive cannabis users (CU)^68^, we examined the clinical application potential of our decoders (4) to detect ER deficits in CU compared to healthy control (HC) participants (n_HC_=48, n_CU_=49) (Fig.1d).

**Fig. 1.**
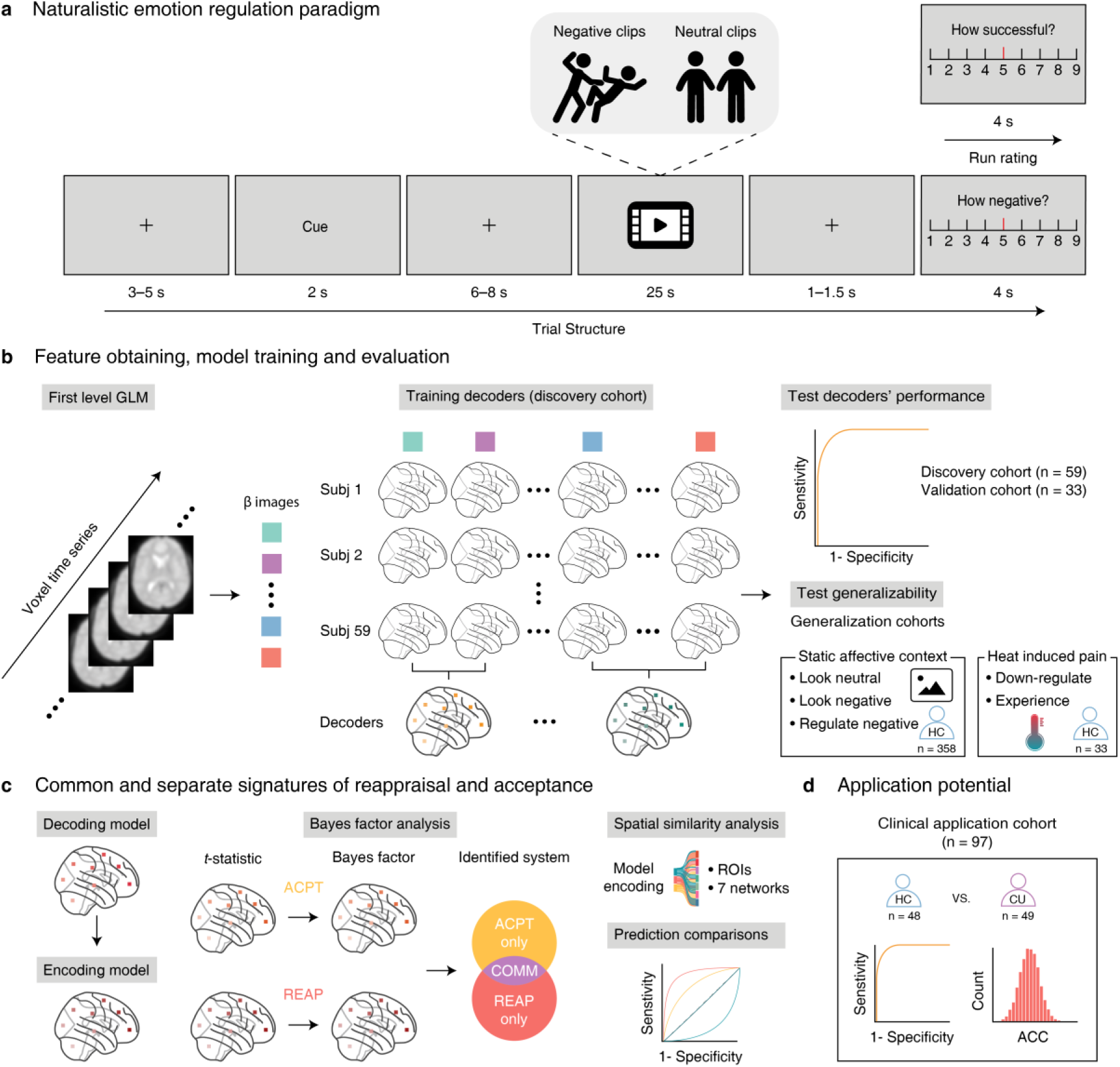
Task paradigm structure, model evaluation, and general analytic workflow. **a,** The naturalistic ER paradigm used in the discovery and validation cohort. The cues were NNES, NERS-A, or NERS-R, indicating which strategy participants should use to view the neutral/negative video clips. At the end of each trial, participants rated their negative feelings. At the end of each run, participants rated their average success level in this run. Notably, the schematics were used only for illustrative purposes to avoid copyright issues and were not part of the original stimulus set. **b**, Feature estimation, model training, and evaluation. First-level contrast maps were used as features in the prediction analysis. The whole brain multivariate pattern predictive of the acceptance, reappraisal, or reaction to the naturalistic stimulus was trained on the discovery sample (n=59) using the support vector machine (SVM) and further evaluated in discovery (cross-validated), validation (n=33) cohorts. The generalizability of the developed signatures across samples, MRI systems, culture, and stimuli (dynamic versus static, n=358 or heat-induced pain, n=33). **c**, Systematically investigate the common and distinct neurobase of acceptance and reappraisal. Determining the contribution of specific brain systems by using univariate and multivariate methods. The performance of isolated brain regions or systems was tested by multiple prediction analyses. **d**, Testing the specificity of developed models in terms of distinguishing corresponding mental processing from other decoders. Subj, subject; NERS-A, naturalistic emotion regulation signature-acceptance; NERS-R, naturalistic emotion regulation signature-reappraisal; NNES, naturalistic negative emotion signature; PINES, picture-induced negative emotion signature from Chang et al.^69^; HC, healthy control; CU, cannabis users.

## Results

### Assessment of the naturalistic ER paradigm

#### Subjective experience and regulation of negative emotions

During fMRI, participants were instructed to react naturally to neutral or negative video clips or use corresponding ER strategies based on presented cues, then report their momentary negative affect on a 9-point Likert scale (9-very negative, 1-not negative at all). In both discovery (n=59) and validation (n=33) cohorts, significant main effects of condition (stimulus type: NV, NeutV; strategies: NV, NA, NR) were observed with repeated-measures analysis of variance (ANOVA) and further simple effects analysis, indicating that the negative clips induced considerable negative emotional experience (discovery cohort: *P*=9.96×10^-39^, 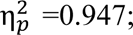 validation cohort: *P*=8.15×10^-17^, 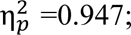 Fig.2, details in supplements) and both strategies allowed participants to effectively attenuate the negative emotions(discovery cohort: *P*=3.18×10^-19^, 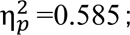 validation cohort: *P*=1.32×10^-12^, 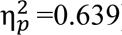). Reappraisal, led to a stronger decrease in negative emotions as compared to acceptance (discovery cohort: *P*=2.02×10^-10^; validation cohort: *P*=2.4×10^-5^). Findings are further corroborated by success rates of ER (discovery cohort: 7.51±1.01, validation cohort: 7.53±0.98).

**Fig. 2.**
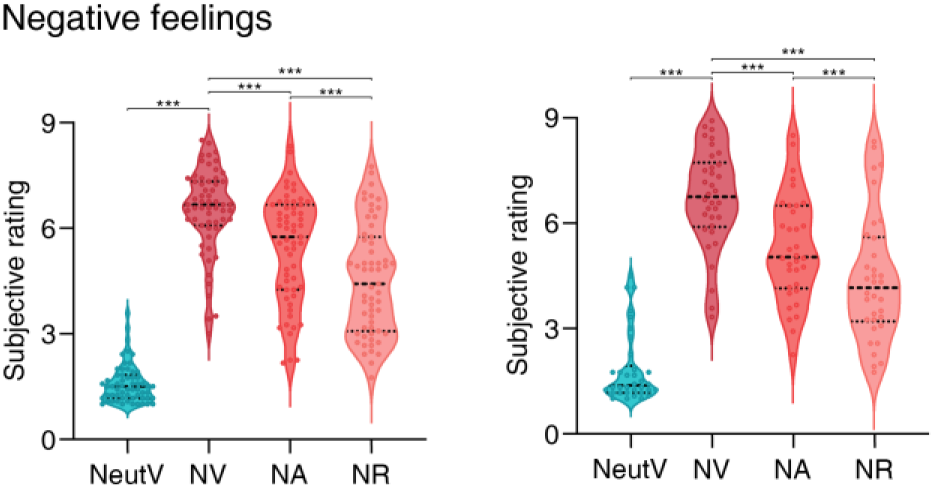
Negative emotional state in the discovery and validation cohort. Averaged subjective feeling ratings in the discovery (left) and validation cohort (right). Violin plots represent the value distribution, each dot represents the negative rating for individual participants averaged across trials in the corresponding condition. The dotted line in the center represents the median and the sides are the quartiles, the range of data points between Q1–1.5IQR and Q3 + 1.5IQR. \*\*\**P*<0.001. NV, view (react to) negative clips; NeutV, view (react to) neutral clips; NA, use acceptance strategy to negative clips; NR, use reappraisal strategy to negative clips.

### Decoding emotional experience and ER strategies in dynamic naturalistic contexts

Employing the support vector machine (SVM) with leave-one-subject-out cross-validation (LOSO-CV) allowed us to identify whole-brain fMRI activation predictive signatures of negative emotional experience (NNES), acceptance (NERS-A), and reappraisal (NERS-R) (Fig.3a, b, and c) during naturalistic emotion processing. Only the data from the discovery cohort were used to develop decoders. The performance of the decoders was validated using cross-validated classification in the discovery cohort and determining the reactivity of the decoders in an independent pre-registered validation cohort (n=33) that underwent a similar paradigm. NNES, NERS-A, and NERS-R could accurately discriminate the corresponding verse control condition, respectively (Fig.3d, Table 1). Testing the signatures with no further model fitting in the validation cohort revealed robust prediction performances in the independent datasets (Fig.3e).

**Fig. 3.**
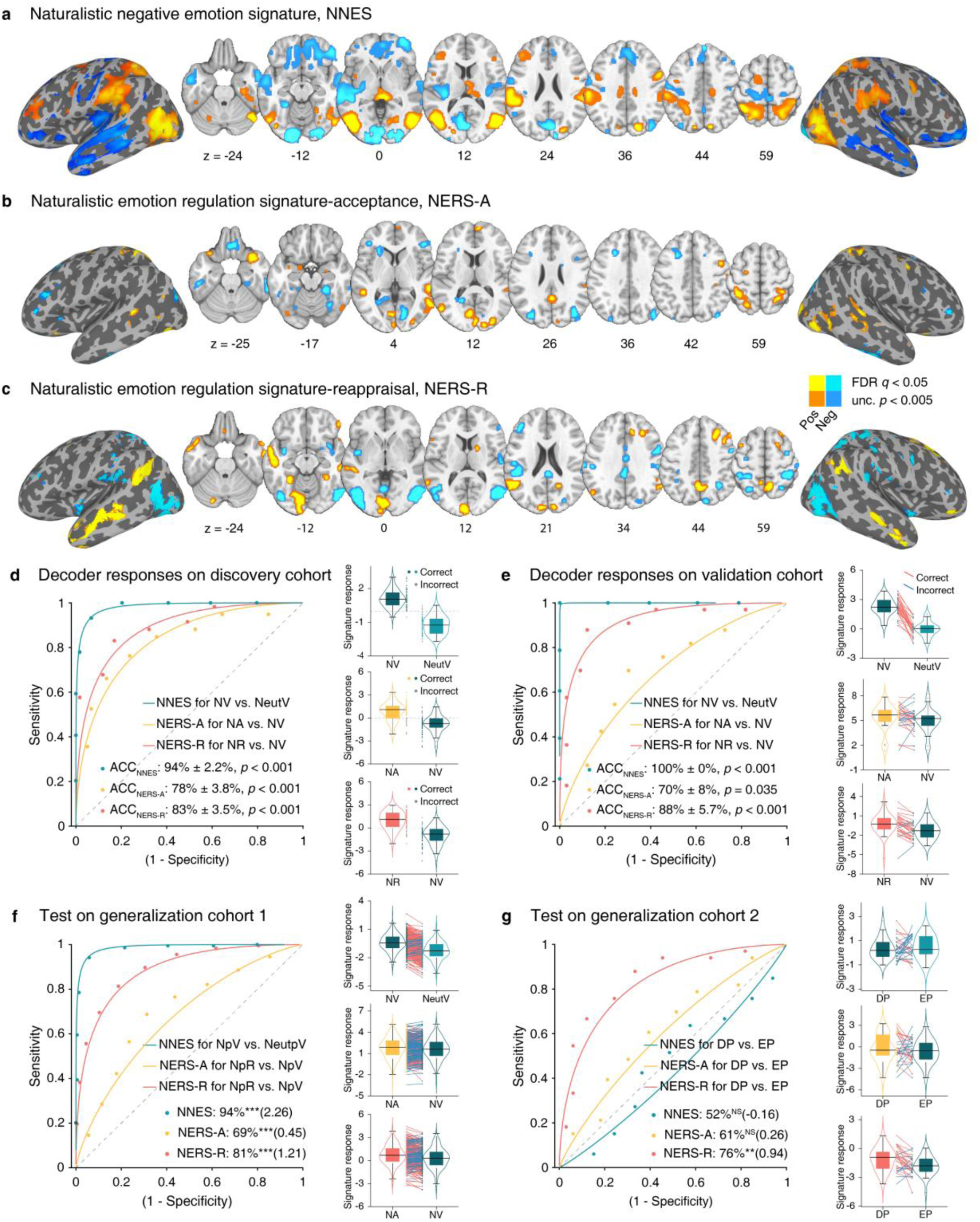
Developed signatures and predictive performances. NNES (thresholded at false discovery rate (FDR) *q*< 0.05) (**a**), NERS-A (FDR *q*<0.05 and unc. *p* <0.005) (**b**), and NERS-R (FDR *q*<0.05 and unc. *p* <0.005) (**c**) pattern maps, based on a 10,000-sample bootstrapping (thresholded at FDR < 0.05). Hot color indicates positive weights, whereas cold color indicates negative weights. **d**, Receiver operating characteristic (ROC) plot illustrates the classification performances of the SVM models on the discovery cohort (n=59), using leave-one-subject-out cross-validation (LOSO-CV) with a misclassification threshold set to 0. The right three panels show the distributions of the decoders’ responses. The boxes are bounded at the first and third quartiles, the whiskers extend to the maximum and minimum values between the median±1.5 quartiles. Data points outside the whisker range are labeled as outliers. The yellow, green, and red dots beside the violin plots indicate correct classification, and the gray dots indicate misclassification. The classification performances of the developed decoders on the validation cohort (**e**, n=33) and generalization cohort (**f**, n=358, **g**, n=33). The red line indicates correct classification, and the blue line indicates misclassification. NNES, naturalistic negative emotion signature; NERS-A, naturalistic emotion regulation signature-acceptance; NERS-R, naturalistic emotion regulation signature-reappraisal; NA, negative (clips)-acceptance; NR, negative (clips)-reappraisal; NV, negative (clips)-view; NeutV, neutral (clips)-view; NpR, negative pictures-reappraisal; NpV, negative pictures-view; NeutpV, neutral pictures-view; ACC, accuracy.

**Table 1.** Classification performance on the discovery and validation cohort.

| Condition | Decoder | ACC (%) |  | Sens (%) |  | Spec (%) |  | Effect | *P* |
| --- | --- | --- | --- | --- | --- | --- | --- | --- | --- |
|  |  |  | SD |  | CI |  | CI | size |  |
| **Discovery cohort (n=59)** |  |  |  |  |  |  |  |  |  |
| NV vs. NeutV | NNES | 94 | 2.2 | 98 | 95–100 | 90 | 81–97 | 3.01 | $<1 \times 10^{-50}$ |
| NA vs. NV | NERS-A | 78 | 3.8 | 75 | 68–86 | 81 | 71–91 | 1.37 | $7.75 \times 10^{-10}$ |
| NR vs. NV | NERS-R | 83 | 3.5 | 83 | 73–92 | 83 | 73–92 | 1.71 | $1.53 \times 10^{-13}$ |
| **Validation cohort (n=33)** |  |  |  |  |  |  |  |  |  |
| NV vs. NeutV | NNES | 100 | 0 | 100 | 100–100 | 100 | 100–100 | 3.55 | $2.33 \times 10^{-10}$ |
| NA vs. NV | NERS-A | 70 | 8 | 70 | 53–86 | 70 | 53–85 | 0.47 | **0.0351** |
| NR vs. NV | NERS-R | 88 | 5.7 | 88 | 75–97 | 88 | 76–97 | 1.42 | **1.10×10<sup>-5</sup>** |
| **Generalization cohort 1 (n=358)** |  |  |  |  |  |  |  |  |  |
| NpV vs. NeutpV | NNES | 94 | 1.2 | 94 | 92–96 | 94 | 92–96 | 2.26 | **<1×10<sup>-50</sup>** |
| NpR vs. NpV | NERS-A | 69 | 2.5 | 69 | 64–74 | 69 | 64–74 | 0.45 | **1.14×10<sup>-12</sup>** |
| NpR vs. NpV | NERS-R | 81 | 2.1 | 81 | 77–85 | 81 | 78–85 | 1.21 | **<1×10<sup>-50</sup>** |
| **Generalization cohort 2 (n=33)** |  |  |  |  |  |  |  |  |  |
| DP vs. EP | NNES | 52 | 8.7 | 52 | 35–69 | 52 | 33–69 | -0.16 | 1 |
| DP vs. EP | NERS-A | 61 | 8.5 | 61 | 44–77 | 61 | 44–77 | 0.26 | 0.2962 |
| DP vs. EP | NERS-R | 76 | 7.5 | 76 | 60–90 | 76 | 60–90 | 0.94 | **0.0045** |

NV, view (react to) negative clips; NeutV, view (react to) neutral clips; NA, use acceptance strategy to negative clips; NR, use reappraisal strategy to negative clips; NNES, naturalistic negative emotion signature; NERS-A, naturalistic emotion regulation signature – acceptance; NERS-R, naturalistic emotion regulation signature – reappraisal; NpV, view negative picture; NeutpV, view neutral picture; NpR, regulate negative emotions induced by picture; DP, Decrease-pain; EP, experience-pain.

### Generalization to independent data

Generalization across samples, MRI systems, culture, and stimuli (dynamic versus static or heat-induced pain) was first determined using data from a large independent cohort (n=358)^28^. NNES could accurately discriminate exposure to negative versus neutral pictures (Fig.3f, Table 1). In line with the strategy used by the participants (acceptance or reappraisal)^28^, NERS-A and NERS-R could accurately classify the ER vs. experience processing.

After excluding the voxels from the visual network^67^(to control the interference of visual processing caused by different materials), NERS-R could accurately distinguish between using the reappraisal strategy to down-regulate vs. experiencing pain induced by heat stimuli, based on the data from the second independent cohort (n=33, Fig.3g). Yet NERS-A and NNES showed poor predictive performance of reappraisal.

Together, these results underscore the robustness and specificity of the decoders for specific mental processes.

### Common and separate signatures of reappraisal, acceptance, and negative emotional experiences

We next systematically evaluated which brain regions provided a consistent and robust contribution to negative affect and were regulated by the respective ER strategies.

#### Bootstrap tests and encoding model estimation

We initially thresholded the multivariate patterns using bootstrapping (Fig.3a, b, and c) and – given that the decoding model features may not only reflect neurobiological processes but also noise components^70^ – next transformed the bootstrapped within-subject weighted maps to reconstructed ‘activation pattern’ (encoding model)^71,72^ and estimated the population-level reconstructed ‘activation pattern’ (FDR *q*<0.05, Fig. 4a).

**Fig. 4.**
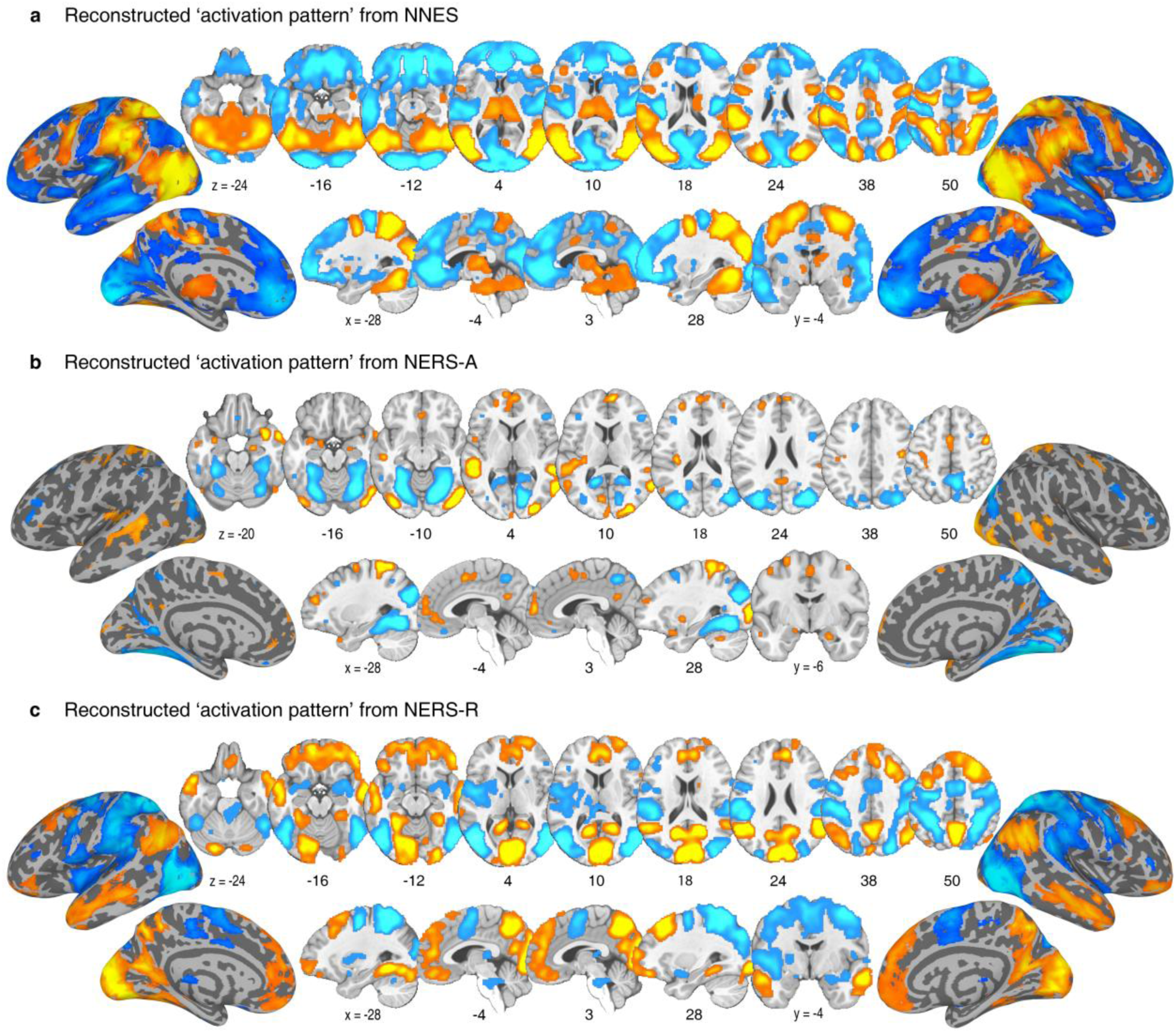
Reconstructed ‘activation pattern’. FDR corrected (*q*<0.05) group-level reconstructed ‘activation pattern’ transformed from NNES (**a**), NERS-A (**b**), and NERS-R (**c**). The color indicates the direction of the relationship between the variable and the model without controlling for other variables – i.e., which voxels are positively (red) or negatively (blue) related to ER strategies. The hot color indicates positive associations, whereas the cold color indicates negative associations.

Aggressive negative emotions induced by watching the negative (versus neutral) clips were accompanied by a positive association with activity in subcortical regions engaged in emotional reactivity, such as the insular, thalamus, periaqueductal grey (PAG), as well as superior parietal (e.g. supramarginal), ventrolateral occipital and posterior mid-temporal regions involved in the integration of information and semantic content^41,73,74^; negative association with activity in a widespread bilateral anterior temporal and frontal network, including the pre-supplementary motor area (SMA) as well as medial and lateral frontal regions.

Compared to experiencing negative emotions, acceptance significantly attenuated activity in subcortical (e.g. fusiform and lingual gyrus), superior and inferior parietal, inferior temporal, and bilateral pars triangularis (IFSa) regions, while concomitantly associated with activity in the amygdala (extended to hippocampus), insular cortex, putamen, frontal regions including the prefrontal lobe (bilateral dorsolateral (dlPFC), dorsomedial (dmPFC), ventromedial (vmPFC), ventrolateral (vlPFC) prefrontal cortex, left rostral anterior cingulate cortex (rACC)), isthmus cingulate, precentral, lateral occipital cortex, SMA, and superior parietal regions including the precuneus (Fig. 4b).

Reappraisal (versus experiencing negative emotion) is negatively associated with activity in subcortical regions, including the bilateral amygdala, insular, putamen, and thalamus involved in emotional experience, as well as lateral occipital, posterior frontal, and parietal regions, including the SMA and supramarginal gyrus. Reappraisal is concomitantly associated with activity in a bilateral frontal network encompassing medial, orbitofrontal, vlPFC, rACC, right dlPFC, as well as parietal regions including the precuneus, superior, and middle temporal regions (Fig. 4c). Univariate analyses additionally confirmed the observed effects (Extended Data Fig.1).

#### Brain systems identified by BF analysis

BF analysis was used to determine the common and separate neural basis between acceptance and reappraisal. ‘Acceptance only’ brain regions were identified by activation during acceptance (BF>5) but not for reappraisal (BF<1/5); reverse for ‘Reappraisal only’ (reappraisal BF>5, acceptance BF<1/5); ‘Common’ brain regions were identified by activation during both acceptance (BF>5) and reappraisal (BF>5)^28,64^.

Effective ER via both strategies robustly recruited cortical midline DMN regions including the bilateral dmPFC (extending to rACC), right inferior precuneus, left cuneus, middle temporal and lateral occipital regions (Fig.5a, see also conjunction analysis in Extended Data Fig.2). Acceptance and reappraisal specifically recruited distributed process-specific systems spanning several cortical systems, with the acceptance primarily engaging somatomotor (and to a lesser extend ventral attention and frontoparietal) large scale networks and subcortical systems (e.g., superior temporal lobe, dlPFC, vlPFC, insular, amygdala, and putamen), while reappraisal primarily engaged an extensive the frontoparietal and DMN regions (e.g., superior frontal and inferior parietal regions, dlPFC, vlPFC, supramarginal, and precuneus).

**Fig. 5.**
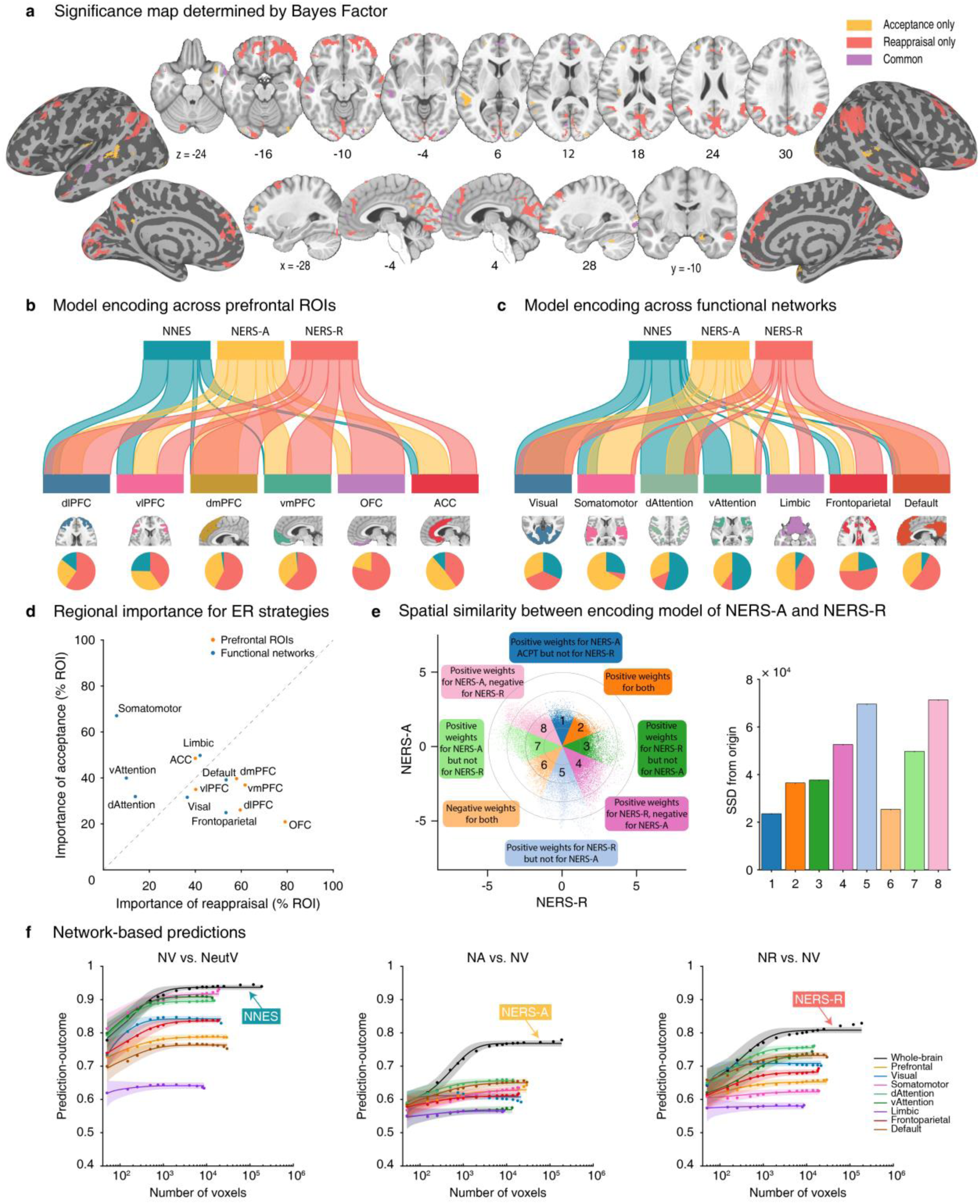
Comparing acceptance, reappraisal, and negative emotion signatures. **a,** Brain regions of the class of each voxel (Acceptance only, Reappraisal only, or Common). River plots illustrated the spatial similarity (cosine similarity) between reconstructed ‘activation pattern’ and anatomical parcellation of the prefrontal cortex^66^ (**b**) as well as resting-state-based functional parcellation of cortical regions^67^ (**c**). Reconstructed ‘Activation patterns’ were FDR q<0.05 thresholded and used positive voxels only for similarity calculation and explanation. In **b** and **c**, the thickness of the ribbons indicates the normalized maximum cosine between sets. The pie charts indicate the relative contributions of each pattern to each ROI (**a**) or network (**b**) (i.e., the percentage of voxels with the highest cosine similarity for each map). **d**, Regional importance scores for acceptance versus reappraisal. X-axis–reappraisal model importance (percentage of ROI or network occupied by the encoding model of reappraisal), y-axis– acceptance model importance (percentage of ROI or network occupied by the encoding model of acceptance). The gray line represents equal importance in both models; a greater distance between the points and the line indicates less equal importance in the two models. The region on the left side has a higher importance for acceptance than for reappraisal, and on the right side is the opposite. **e**, Voxel-level spatial similarity between the encoding model of NERS-A and NERS-R. Scatter plots illustrate normalized unthresholded voxel beta weights of the reconstructed ‘activation pattern’ of NERS-A (y-axis) and NERS-R (x-axis). Colored octants indicate voxels of shared positive (Octants 2) or shared negative (Octants 6), selective positive weights for NERS-A (Octant 1) or NERS-R (Octant 3), selective negative weights for NERS- A (Octant 5) or NERS-R (Octant 7), and voxel weights opposite for the two signatures (Octants 4 and 8). The right bars indicate the sum of squared distances from the origin (0, 0) for each octant, which integrates the number of voxels and combined weights. **f**, Regulating or experiencing naturalistic negative emotions is distributed across multiple systems. The model performance was evaluated as increasing numbers of voxels/features (*x*-axis) were selected to predict experience naturalistic negative emotions (NV vs. NeutV), use acceptance (NA vs. NV), and reappraisal (NR vs. NV) strategy in different ROIs including the whole brain (black), prefrontal cortex (light orange), or individual large-scale cerebral networks (other colored lines). The *y*-axis denotes the cross-validated classification accuracy. Colored dots indicate the mean correlation coefficients, solid lines indicate the mean parametric fit, and shaded regions indicate the s.d. dlPFC, dorsolateral prefrontal cortex; vlPFC, ventrolateral prefrontal cortex; dmPFC, dorsomedial prefrontal cortex; vmPFC, ventromedial prefrontal cortex; OFC, orbitofrontal cortex; ACC, anterior cingulate cortex; vAttention, ventral attention; dAttention, dorsal attention.

#### Spatial similarity between stable decoding maps and interest networks, as well as crucial ROIs

We further estimated the spatial similarities between the reconstructed ‘activation pattern’ and prefrontal anatomical regions consistently involved in emotional experience and regulation (ROIs)^66^ as well as large-scale functional networks^67^ (Fig.5b, c). For each region, we calculated the ratio of importance for acceptance versus reappraisal versus experience negative affect (pie plot in Fig.5b, c; Supplementary Table 1), with importance defined as the percentage of the region encoded by each model^42,75^, while also directly comparing acceptance and reappraisal (Fig.5d). Except the vlPFC, which was engaged in all processes, prefrontal regions exhibited a stronger engagement during both regulation strategies as compared to experiencing the emotions. While the vmPFC, dmPFC, and vmPFC showed comparable contributions to both strategies, the ACC had stronger engagement for acceptance, whereas the dlPFC and orbitofrontal cortex (OFC) exhibited stronger engagement during reappraisal. On the network level, the visual networks exhibited equal importance across the three models, the experience of negative emotion may mostly encoded in the dorsal, ventral attention, and somatomotor networks. When comparing between ER strategies, the somatomotor and attention networks presented higher encoding importance for acceptance, while the frontoparietal networks had relative stronger contribution for reappraisal and the DMN exhibited similar importance for both strategies.

To provide more comparative pictures of how acceptance and reappraisal are encoded, we analyzed the voxel-level spatial covariation between the unthresholded normalized (z-scored) encoding maps of NERS-A and NERS-R^76^. Fig.5e illustrates the joint distribution of the voxel weights of the encoding model of NERS-A and NERS-R. Octants with different colors indicate voxels of shared positive/negative, selective positive/negative weights, and voxel weights opposite for the two signatures. Stronger weights (sum of squared distances to the origin (SSDO)) were shown in the nonshared octants (5,7) as well as opposite octants (4,8), and fewer weights in the shared octant 6, further supporting that separable brain representations contribute to implementing acceptance and reappraisal strategies.

#### Predictive performances between decoders

To determine whether our decoders reflected both the common and separate processing of naturalistic acceptance, reappraisal, and negative emotion, we also compare the decoders’ performances on different conditions from the validation cohort and add a neural signature for negative affect (PINES, picture-induced negative emotion signature^69^). As shown in Extended Data Fig.2c, except for NERS-R, the other three decoders could distinguish between NV and NeutV, but NNES has the highest accuracy and effect size. NNES and PINES show low performance when predicting ER processing (acceptance and reappraisal). Only NERS-A could accurately classify between NA and NV. Both NERS-R and NERS-A could classify between NR and NV, but NERS-R is more accurate with a larger effect size. Together, these findings indicate that our decoders captured and further demonstrated the common and separate nature of the above three mental processes.

#### Evaluation of predictive performances of local systems

Given the continuing debate about the specific systems underlying ER, including the dlPFC^27,28,77^, and the DMN^34,78,79^, we (1) employed network-based and searchlight-based analyses to estimate the predictive performances of local brain regions and (2) compared their performances with whole-brain decoders. To control for the potential effects of the number of voxels/features in prediction (i.e., the whole-brain model contains many more features), voxels were randomly selected (repeated 1,000 times) from a uniform distribution spanning the whole brain (Fig.6b, black), prefrontal cortex (light orange, which has been suggested play crucial roles in both generate and regulate emotions^26,27,31^), or individual large-scale functional networks (averaged over 1,000 iterations)^41,42,80^. The asymptotic prediction when sampling from the whole-brain, as we trained decoders (black line in Fig.6b), was more accurate than the asymptotic prediction within individual networks (colored lines), especially with more available voxels (details in Supplementary). Moreover, model performance was optimized (i.e., reaching asymptote) when approximately 10,000 voxels were randomly sampled across the whole brain, further confirming that information about ER processing is contained in patterns of activity that span multiple systems. The above results can also be demonstrated by searchlight analysis, although statistically significant and thresholded brain regions are similar to the whole-brain decoders (Extended Data Fig.3), the effect sizes in terms of prediction–outcome accuracy were substantially smaller than those obtained from the developed decoders (*P*<0.001). Together, findings underscore that ER and negative emotional experiences are encoded in distributed neural patterns that span multiple systems, rather than single brain regions or networks.

### Translation into a neuromarker for determining ER deficits in MDs

To test whether our decoders can capture deficient ER processing in individuals with MDs^52,68,81^, we applied the decoders to extended data from our previous study^68^ describing ER deficits in cannabis users (CU). For the present study, we incorporated additional unpublished data from individuals with CU disorder and matched controls who underwent a similar picture-based reappraisal paradigm with the concomitant fMRI system (details see ref.^68^, study protocol NCT02801214, clinical-trials.gov).

A total of 48 HC participants and 49 CU were included in the extended dataset. In line with the findings in the initial study, behavioral analyses confirmed ER deficits in the CU (reappraisal success^68^, 0.46±0.46) as compared to HC (0.78±0.54) (*t*=-3.072, *p*=0.003, Cohen’s d=0.50) in the extended sample.

NERS-R could accurately predict neural activity during cognitive reappraisal (distancing) (NpD) vs. view (NpV) negative pictures in HC participants but not in CU (Fig.6, Table 2). NNES could accurately distinguish between NpD and view neutral pictures (NeupV) in both groups, underscoring impaired regulation rather than excessive emotion reactivity as a neurofunctional marker for CU. Further permutation tests revealed that the pattern expression of the two groups has significant differences for NERS-R and NNES (both *P*<0.0001, Fig.6). In contrast, NERS-A showed poor predictive performance.

**Fig. 6.**
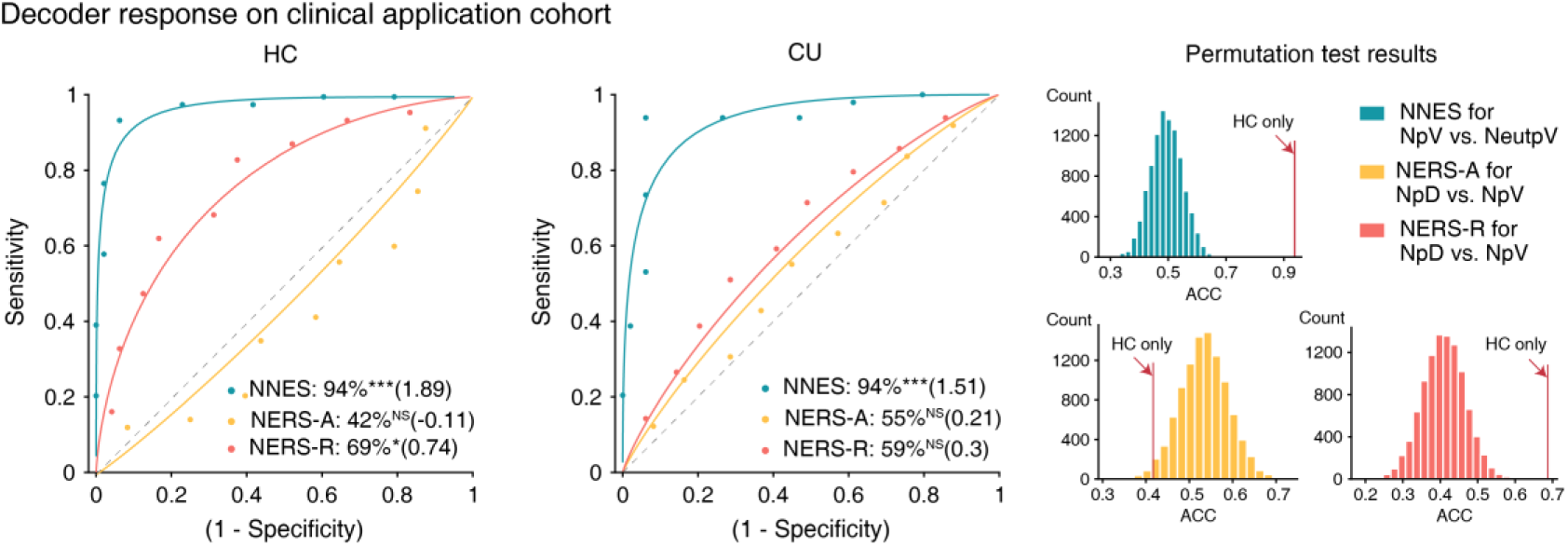
Testing application potential. The NERS-R could accurately distinguish between distancing (NpD, another form of reappraisal) and viewing (NpV) negative pictures in health control (HC, n=48) participants but not for cannabis users (CU, n=49). Permutation tests reveal that the pattern expression of the two groups has significant differences (red bar chart). NNES could accurately distinguish between NpV and view neutral pictures (NeutpV); the pattern expressions of the two groups are also significantly different (green bar chart). \**P*<0.05, \*\*\**P*<0.001, NS not significant (binomial test, two-sided, uncorrected).

**Table 2.**
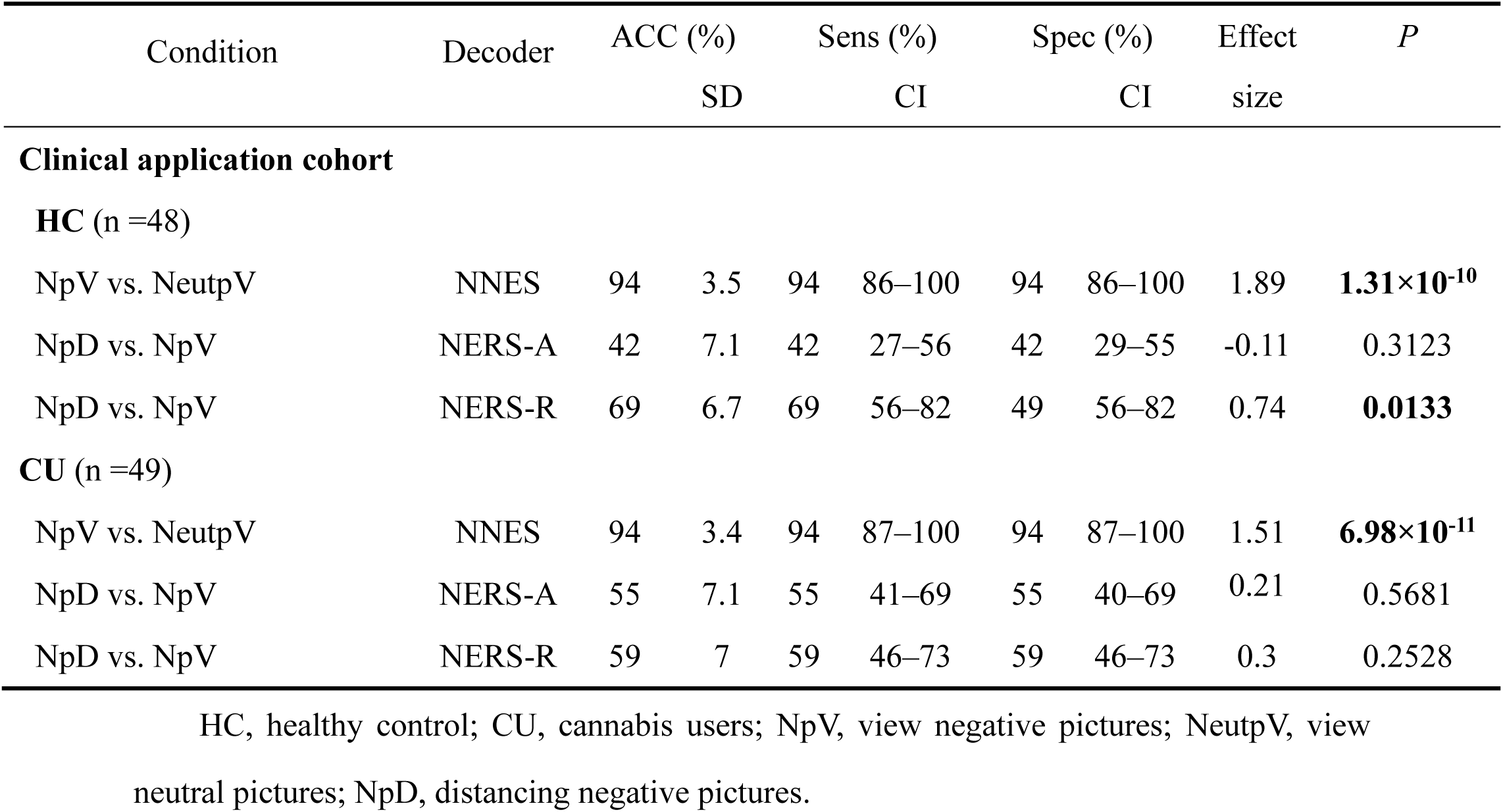
Classification performance on the clinical application cohort.

## Discussion

Identifying comprehensive and accurate brain models for the common and distinct brain representations that underly ER is critical for advancing mechanistic models of cognitive regulation as well as for developing accurate biomarkers for regulatory dysfunction in MDs. While reappraisal and acceptance are widely implemented in therapeutic contexts and have been debated from theoretical^1,3^, experimental^28^, and clinical^12,82^ perspectives, the extent to which they rely on common and dissociable systems in ecologically valid dynamic contexts remains unresolved.

Here, we combined immersive naturalistic neuroimaging with predictive modeling and BF analyses to systematically establish and validate three comprehensive, accurate, and sensitive brain-based models for negative affect (NNES) and its regulation via acceptance (NERS-A) and reappraisal (NERS-R). The developed decoders are generalizable across imaging systems, cultures, and paradigms. Next, through a series of systematic analyses, we uncovered the shared and separable neural representations of acceptance, reappraisal, and negative emotion, which indicated that these mental processes are encoded in distributed and distinguishable neural systems on the whole brain level, yet also show process-specific engagement of large-scale systems. Both acceptance and reappraisal engaged cortical midline regions of the core DMN, including the dmPFC and posterior parietal cortex, indicating a general contribution of this system to effective ER. Each strategy, furthermore, exhibited distinct profiles aligned with its cognitive demands. Acceptance was encoded in distributed regions spanning subcortical, somatomotor, and ventral attention networks, consistent with embodied awareness and experiential processing. Reappraisal, by contrast, relied on regions spanning the fronto-parietal control network, including lateral frontal and inferior parietal regions, reflecting its reliance on executive functions and semantic updating. While the distinction was further substantiated by BF analyses and spatial similarity mapping, decoding performance was substantially higher for whole-brain decoders as compared to regional models, underscoring the importance of brain-wide integration for capturing emotional experience and regulation. Finally, the neurofunctional decoders could detect reappraisal-specific ER deficits in a large set of CU, underscoring the sensitivity of the signatures to detect specific behavioral impairments on the neural level and clinical potential.

Combining naturalistic fMRI with mass-univariate and multivariate predictive analyses, we established comprehensive and distributed brain models for negative emotional experiences and their cognitive regulation in dynamically close-to-real-life contexts. The affective videos induced immersive and strongly negative emotional states, which were accompanied by positive engagement of brain regions involved in emotional experience, physiological and affective arousal, including the insula, PAG, thalamus^26,27,74,75,83^, and negatively associated with the engagement of a widespread frontal and temporal network involved in cognitive and regulatory control. Spatial similarity and network-level analyses further confirmed the contribution of large-scale cortical networks, including the attention and visual networks (resembling previous studies^84^ using dynamic stimuli). Importantly, both the developed (NNES) and an established signature for negative affect (PINES) poorly predict the ER strategies, supporting previous research indicating that emotion generation and regulation are neurally separable processes^25–27^.

Both ER strategies could efficiently reduce negative emotional states, indicating that utilizing reappraisal and acceptance represents an efficient means to regulate negative emotions and the associated neural systems in dynamic, ecologically-valid contexts. In line with ref.^22^, reappraisal led to a greater reduction of subjective negativity compared to acceptance.

On the neural level, ER via both strategies was predicted by neural regulation in distributed cortical and subcortical systems, with common and distinct signatures. BF and conjunction analyses demonstrated that effective ER by both strategies engaged core regions of the DMN that are strongly involved in the evaluation of the mental state of oneself and others (dmPFC)^19,26,31^, episodic autobiographical memory (inferior precuneus)^85,86^, rapid emotional conflict adaptation (rACC)^87^, self-awareness and interpersonal processing (temporal regions)^88,89^. Given that both ER strategies require the appraisal of the emotional state concerning the self, the dmPFC and other core DMN regions may support appraisal, metacognitive evaluation, and internal self-reference, enabling ER based on internal goals^90^. On the anatomical level, these functions of the DMN, in particular the dmPFC, may be supported by interconnections with the rostromedial prefrontal, lateral orbitofrontal (lOFC), and pregenual ACC, which may support its role as a central hub for the implementation of both ER strategies^31,90^. The effective regulation was accompanied by reduced engagement of a widespread bilateral network including bilateral superior parietal, inferior and precentral frontal, and lateral occipital regions, which may reflect attenuated motor readiness and expressive output, as well as attenuated visual processing of emotionally salient stimuli, and in turn may promote regulatory control across both strategies^91–93^.

Distinguishable contributions were observed in somatomotor^94^, attention, or the frontoparietal control network^79,95^, respectively. In line with the notion that reappraisal represents an active and effortful ER strategy, effective implementation specifically engaged an extensive bilateral network of distributed regions within the fronto-parietal control network. These included lateral frontal regions involved in working memory (dlPFC)^77^, general and valence-specific inhibition (vlPFC, lOFC)^26,31,96,97^, and medial prefrontal regions involved in computation of value or safety (vmPFC, mOFC)^31,98–100^. Parietal regions were specifically recruited during reappraisal, including regions involved in self-referential processing (superior precuneus)^101–103^, attentional control (inferior precuneus)^104^, and conscious representation of affective semantic content (e.g., Griffiths et al.,^105^). In contrast, reappraisal specifically decreased engagement of brain regions traditionally linked to emotional “reactivity”, including subcortical systems such as the amygdala^26,27,98,104,106–109^ and the anterior insular cortex^98^.

Acceptance was also encoded in some distinguishable but rather focal prefrontal regions (e.g., dlPFC), yet showed stronger engagement of distributed regions spanning the somatomotor (e.g., superior parietal lobe, SMA, posterior insula)^110,111^ and ventral attention networks (e.g., preSMA). Compared to reappraisal, enhanced engagement of these regions may reflect that effective acceptance relies on goal-oriented attentional deployment (superior parietal lobe^53,112,113^), interoception (posterior insula^114,115^), and embodied cognition-affect processing (SMA/preSMA^44^). Notably, acceptance is specifically negatively associated with the brain systems involved in self-referential processing (superior precuneus)^101–103^ and threat perception (fusiform)^116,117^, while it was positively associated with activation in subcortical systems traditionally linked to emotional reactivity, including the amygdala, hippocampus, and putamen. The stronger engagement of these systems may reflect the non-judgmental awareness of affective arousing and sensory experience and may promote hippocampus-dependent encoding and contextualization^118^, in turn facilitating adaptive integration of the emotional experience with autobiographical and situational contexts^22,118–121^.

Comparative predictive analyses using the whole-brain decoders further substantiated common yet, on the whole-brain level, also clearly distinguishable neural representations, such that the decoders exhibited a considerably higher accuracy for the targeted ER strategy. The importance of individual brain regions or networks in the experience and regulation of negative emotions has been emphasized in previous frameworks^25–27^. While our findings support preferential engagement of specific networks by distinct strategies – the fronto-parietal control network during reappraisal and the somatomotor network during acceptance - our large-scale network- and searchlight-based predictive analyses further highlight that, under dynamic and naturalistic conditions, both strategies rely on the coordinated activity of distributed and distinguishable brain-wide representations. The distributed engagement was further supported by a system-specific analysis of the prefrontal cortex, commonly identified as systems promoting distinct ER strategies^25–27^. While most prefrontal systems contributed to both strategies, the orbitofrontal cortex showed a markedly higher engagement during reappraisal, which may be due to its extensive anatomical connections and functional coupling with the amygdala during reappraisal, and which is underscored as this region is critical for mediating effective reappraisal of negative emotions^92,122–124^. In general, the distributed signature of the whole-brain model showed the highest predictive performance, which is in line with decoders for other mental processes, e.g., digust^42^, fear^41^, or pain^80^.

Importantly, our neurofunctional signatures for ER demonstrated a clinical translational potential, such that the decoders accurately tracked the ER strategy-specific neural representation of impaired reappraisal success. The NERS-R showed an accurate reappraisal prediction in HC but failed to distinguish reappraisal from negative experience in individuals with CU and reappraisal deficits on the behavioral level, with permutation tests indicating that NERS-R and NNES could capture the abnormal brain activation pattern among CU. Impaired ER and altered emotion reactivity have been widely demonstrated across multiple MDs, including CU^68,125,126^, opioid use disorders^81,127,128^, alcohol use disorders^129,130^, cocaine use disorder^131,132^, major depressive disorder^133,134^, anxiety disorders^135,136^, yet precise neuromarkers to track emotional deficits on the neurofunctional level are lacking. Given the critical and transdiagnostic role of ER deficits in MDs, the present multivariate predictive patterns may facilitate the development of brain-based diagnostic and treatment response markers. Our marker accurately tracks emotional reactivity and its effective modulation via cognitive strategies, the marker may also serve as a target state for behavioral as well as brain-based intervention (e.g.,^108,137^). Further studies are required to determine common and mental disorder-specific alterations in these markers to validate the decoders’ accuracy, clinical utility, and sensitivity.

Limitations of the present study are inherent to technical limitations. While combining fMRI with MVPA gives a relatively precise way to investigate brain activity, and has been revealed to be consistent with the conventional univariate approach, limitations are inherent to fMRI (e.g., indirectly measuring neuronal activity, 3T fMRI may not be able to precisely localize to the midbrain and brainstem nuclei^75^), and more precise measurement may support our understanding of the relationship between ER and brain activation.

In conclusion, we developed and validated three robust, generalizable, and clinically translatable whole-brain neural signatures for negative emotional experience and its effective regulatory control via acceptance and reappraisal in dynamic naturalistic contexts. Our within-subject design enabled a systematic comparison of the common and distinct neural bases underlying these ER strategies and suggests that core DMN regions mediate effective regulation across strategies. On the whole brain level, the strategies were distinguishable, and the predictive neurofunctional signatures demonstrated high robustness across independent cohorts, experimental paradigms, and MRI systems. The findings emphasize that an effective ER is encoded not in isolated brain regions but in synergistic, distributed whole-brain activity patterns.

Application to individuals with ER deficits underscored that the neural decoders offer substantial translational potential. Beyond accurately detecting ER deficits in individuals with MDs, future applications include: (a) serving as neural markers to assess intervention effects targeting acceptance and reappraisal; (b) characterizing ER-specific neural impairments across MDs; (c) delineating common and distinct neural architectures of various ER strategies; and (d) informing frameworks centered on the role of the DMN in ER. Together, this work provides a foundation for developing mechanistically-informed, brain-based targets and outcome measures for clinical interventions aimed at enhancing emotional resilience.

## Methods

### Participants

#### Discovery cohort

Sixty-eight healthy participants were recruited from local universities. Exclusion criteria included left-handed, color blindness or weakness, and current physical or mental illnesses. Six participants were excluded due to excessive head motion (> 3 mm or 3°), and three due to failed post-check. Fifty-nine participants (19 male, 21.3±1.7 years old, mean±SD) were included in the final analysis.

#### Validation cohort

Thirty-four healthy participants were recruited as the validation cohort (11 male, 20.3±1.4 years old, mean±SD), with the same exclusion criteria as the discovery cohort. One participant was excluded due to being detected as an outlier by the fmri_data.mahal function in the Canlabcore toolbox (*P*<0.05 Bonferroni corrected).

All participants provided written informed consent. All participants were compensated 140 RMB after the experiment. This pre-registered study (discovery cohort: https://osf.io/s3bp2, validation cohort: https://osf.io/68jkw) was approved by the ethics committee of the University of Electronic Science and Technology of China and in accordance with the Declaration of Helsinki.

### Stimuli and Paradigm

#### Discovery cohort

A total of forty-eight silent video clips were used in the current ER paradigm, rated by one presented once by E-prime software (Version 3.0; Psychology Software Tools, Sharpsburg, PA). Twelve neutral clips include normal driving records, surveillance, vlogs, etc.; thirty-six negative clips include threatening animals, human fights, bullying, car accidents, horror movies, etc. Neutral and negative clips are matched by basic scenarios and characters.

The experiments consisted of four conditions: react (view)-neutral (NeutV), react-negative (NV), reappraise-negative (NR), and accept-negative (NA), each with twelve clips, divided into four runs. To alleviate the potential cognitive overload caused by switching mindsets too often or the potential fatigue or boredom caused by using the same mindset for a long period, we combined “react” with “reappraisal” or “react” with “accept” in each run, and use ABBA method between participants to balance order effect (see Fig.1a).

Participants were provided with written instructions (see supplements), asked to use a certain way of thinking (“react”, “reappraisal”, or “accept”) to subsequently video clip, and received ∼20 mins training before scanning. Each trial began with a fixation cross (3–5s), followed by a 2s cue, then a fixation cross last 6–8s, after that a 25s video clip, followed by a fixation cross (1–1.5s), then a 9-point Likert scale (4s) for participants to report “How negative do you feel right now?” (9 - very negative, 1 - not negative at all). At the end of each run, participants will rate “On average, how successfully have you used the indicated mindset in this run?” (9 - very successful, 1 - not successful at all).

#### Validation cohort

The stimuli, experiment conditions, and trial structure are identical to the discovery cohort. However, to control the potential influence of the “ER trial” on the “react trial” in the same run, we placed all 12 trials of each condition within one run (randomly presented), using the Latin square design to balance order effects between participants.

### MRI data acquisition and preprocessing

#### Discovery cohort

MRI data were collected on a 3.0 T scanner (Ingenia Elition; Philips Healthcare, Best, Netherlands) with a 32-channel head coil. Structural images were acquired using high-resolution three-dimensional T1-weighted images (repetition time (TR)=8.2 ms, echo time (TE)=3.8 ms, field of view (FOV)=243 mm, flip angle (FA)=8°, 243×243 matrix, 1×1×1 mm voxels, 180 sagittal slices, phase encoding anterior » posterior) and were used for anatomical localization and warping to the standard Montreal Neurological Institute (MNI) space only. Functional images were acquired with an echo planar imaging-free induction decay (EPI-FID) sequence (TR=2000 ms, TE=30 ms, FOV=240 mm, FA=80°, 80×80 matrix, 3×3×3.2 mm voxels, 34 interleaved ascending axial slices, phase encoding posterior » anterior). In total, four runs of each 268 measurements were acquired. Preprocessed was carried out using the Statistical Parametric Mapping (SPM 12, https://www.fil.ion.ucl.ac.uk/spm). The first five volumes of each run were removed, the different acquisition timing of each slice was corrected and realigned to the first volume, and nonlinear distortions related to the head motion were corrected by unwarping. The high-resolution anatomical image was segmented and co-registered with the functional images to the generated skull-stripped structural image. The functional images were normalized to Montreal Neurological Institute (MNI) space, interpolated to 2×2×2 mm^3^ voxel size, smoothed by 8-mm full-width at half maximum (FWHM) Gaussian kernel, and bias field corrected. Image intensity outliers were identified based on meeting any of the following criteria: (a) signal intensity >3 standard deviations from the global mean, (b) signal intensity and Mahalanobis distances >10 mean absolute deviations based on moving averages with a full-width at half maximum (FWHM) of 20 image kernels. The time points marked as outliers were severed as separate nuisance covariates included in the first-level model. Additionally, the design matrix included negative feeling ratings (1−9) as a parametric modulator for the reaction or regulation period.

#### Validation cohort

MRI data were collected on a 3.0 T scanner (Vida; Siemens Healthineers, Erlangen, Germany) with a 64-channel head coil. Structural images were acquired using high-resolution T1-weighted images by the original three-dimensional Single-shot TurboFLASH sequence from Siemens (TR=2300 ms, TE=2.32 ms, FOV=240 mm, FA=8°, 256×256 matrix, 0.9×0.9×0.9 mm voxels, 192 sagittal slices, phase encoding posterior » anterior) and were used for anatomical localization and warping to the standard Montreal Neurological Institute (MNI) space only. Functional images were acquired with an echo planar imaging-free induction decay (EPI-FID) sequence (TR=2000 ms, TE=29 ms, FOV=240 mm, FA=90°, 80×80 matrix, 3×3×3 mm voxels, 36 interleaved ascending axial slices, phase encoding posterior » anterior). In total, four runs of each 268 measurements were acquired. Using an identical pre-processing procedure to the discovery cohort (only modifying the parameters based on the sequence setting).

### Mass-univariate analysis

#### Discovery cohort

Subject-level general linear model (GLM) analysis was conducted using SPM12. Twenty-four head motion parameters and indicator vectors of outlier time points were modeled as non-interested regressors ^41^. The fixation-cross epochs served as the implicit baseline, the periods of cue presentation and rating were modeled to eliminate the extra effects on the baseline, and the clips presented period were used as the interest regressor. A high-pass filter of 180s was applied. Additionally, the subjective negativity ratings (1−9) for each clip were included as a parametric modulator for the reaction and regulation period. The primarily interesting contrasts are NR vs. NV, NA vs. NV, and NV vs. NeutV. To avoid the possibility that participants may subconsciously use the corresponding ER strategy at different runs, which may lead to confounding effects of another ER strategy when comparing NA or NR with NV, we modeled NV in the runs with NA and NR, respectively. The contrast beta images were submitted separately to the group-level analysis and used to perform MVPA analysis.

We also conducted a single-trial analysis for MVPA analysis by specifying a GLM design matrix with separate interest regressors for each clip. Nuisance regressors, implicit baseline, and high-pass filters were identical to the above analysis.

#### Validation cohort

The processing procedure is identical to the discovery cohort, except that the NV condition was modeled as a whole since all NV trials were in one run for the paradigm of the validation cohort.

### Multivariate pattern analysis

Following the previous framework^138^, to obtain the robust brain activity patterns that can distinguish and predict between different contrasts mentioned in the mass-univariate analysis section, we trained the SVM classification (kernel linear C=1) decoders by applying LOSO-CV methods using subject-level whole-brain contrast images data (gray matter masked^41^). Of note, only the data from the discovery cohort was used to develop the decoders. To assess the cross-validated performance of the decoders, we calculated the predictive accuracy, sensitivity, and specificity by comparing the prediction and true outcome (threshold=0). Additionally, to test the generalizability of the developed decoders, we applied them to other independent datasets, e.g., the validation cohort, and generalization cohort (see ‘Generalization cohort’ section below) to test the pattern response for each map and the classification capacity.

### Test generalizability with independent datasets

One recent study^28^ explored the ER process and provided subject-level univariate-beta maps from the Adult Health and Behavior project–phase 2 and the Pittsburgh Imaging Project^139^. Briefly, 358 participants performed ER tasks with picture material. Subject-level Reappraisal and acceptance strategies are reported to be used during the scanning^28^. To test decoders’ generalizability, NERS-A, and NERS-R were applied to predict ‘Regulate negative’ vs. ‘Look negative’ and NNES to predict ‘Look negative’ vs. ‘Look neutral’.

We also test whether developed decoders could predict using the reappraisal strategy to “decrease” vs. “experience” the pain induced by heat stimuli, based on the data from the generalization cohort 2 (n=33)^63^. To control the interference of visual processing caused by different materials, we excluded the voxels from the visual network^67^.

### Identifying the neural basis of acceptance and reappraisal, as well as negative emotion response

#### Bootstrap tests and encoding model estimate

To identify robust significant features, we first bootstrapped the discovery cohort of 10,000 samples (with replacement), calculated the two-tailed uncorrected P-value of the predictive weights, and used the False Discovery Rate (FDR) correction to control multiple comparisons (set FDR q<0.05). Given that the signatures of the multivariate backward/decoding model may capture the interested brain activation (e.g., ER) as well as the noise components in the data^70^. For interpretation, we transformed the bootstrapped within-subject pattern to ‘activation patterns’ (forward/encoding model), which is similar to the ‘structure coefficients’, and could indicate the direction of the relationship between the variable and the model without controlling for other variables – i.e., which voxels are positively or negatively related to ER strategies. The group-level significant brain regions were obtained by conducting a one-sample *t*-test (FDR *q*<0.05).

#### Bayes Factor analysis

To identify the brain regions that are acceptance or reappraisal specific or the common regions of those two strategies, we utilized the BF approach to quantify evidence for alternative and null hypotheses^28,64^. Specifically, the BF value of a given voxel reflects the likelihood of the alternative and null (e.g., the neural activity is correlated with naturalistic acceptance or not). The BF values are calculated from *t*-values of a one-sample *t*-test of encoding maps from the discovery cohort, using a Jeffreys-Zellner-Siow (JZS) prior and formula (see below) provided by a previous study^140^.

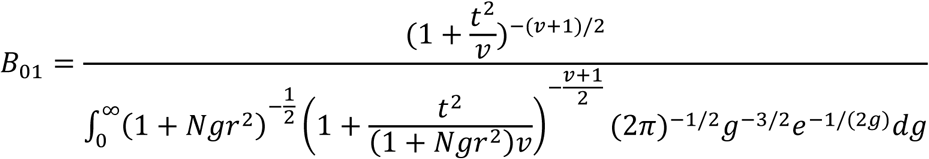

where *N* represents the sample size, *t* for the *t*-statistic*, v* denotes the degrees of freedom, and *r* is the scale factor. In the current study, r was set as 0.707, which suggested a moderate effect size^28,140^.

Because the null is bound by t=0 and the current sample size (N=59), the BF cannot be smaller than .14 but can be infinitely large, a heuristic threshold of 5 was set for the alternative hypothesis and 1/5 for the null to attain moderate to strong evidence in favor of both hypotheses^64,141^. Note that in current datasets, the *P* value threshold of BF=5 is *P*=0.0067, which is smaller than the *q*<0.05 FDR-corrected threshold for the encoding model of NERS-R (*P*=0.0190) but not for NERS-A (*P*=0.0051), hence we further bound the alternative results by the corresponding FDR-corrected threshold, specifically as follows:

- ‘Acceptance only’ voxels – positive activation with BF>5 and P<0.0051 for acceptance, and BF<1/5 for reappraisal.
- ‘Common’ voxels – positive activation with BF>5 and P<0.0051 for acceptance, and with BF>5 and P<0.0190 for reappraisal.
- ‘Reappraisal only’ voxels – positive activation with BF>5 and P<0.0051 for reappraisal, and BF<1/5 for acceptance.

To verify the results of BF analysis, we also performed the conjunction analysis between the thresholded (FDR q<0.05) activation maps of those two strategies obtained from mass-univariate analysis and the reconstructed “activation pattern”, separately.

#### Spatial similarity between stable decoding maps and interest networks, as well as crucial ROIs

The spatial similarity between stable encoding maps and seven large-scale resting-state networks, as well as crucial ROIs (dlPFC, vmPFC, dmPFC, and vlPFC), was illustrated by river plots. The spatial similarity was computed as cosine similarity between the networks or ROIs and the threshold NERS-A, NERS-R, and NNES (FDR q<0.05, positive values only). To further compare the underlying neural representations between NERS-A and NERS-R, we examined the voxel-level spatial covariation between the unthresholded weight maps for those two models (z-scored).

#### Comparing the performance of developed models with PINES

To investigate whether developed decoders capture both the common and separate aspects of naturalistic acceptance, reappraisal, and reaction to negative emotion, we compared the decoders’ performances on data of different conditions from the validation cohort and along with a decoder provided by a previous study^69^ – PINES – which have been demonstrated could track general negative emotion experiences induced by pictures.

#### Evaluation of predictive performances of local systems

To investigate the contribution of a single resting-sate brain network (e.g., DMN) or individual regions to the ER or react processing, we first estimated the prediction performance of seven large-scale networks as well as the prefrontal cortex, which has been demonstrated to have crucial roles in both generate and regulate emotions^26,27,31^ and compared the performance with developed models. Additionally, we employed a whole-brain searchlight-based analysis (three-voxel radius spheres) to test the predictive performance of individual regions.

Together, we combined multiple analytic approaches to systematically interrogate the role of various brain regions or systems in acceptance and reappraisal.

### Clinical application potential

ER and reaction abnormalities have been demonstrated in MDs, e.g., SUDs^52,68,81,126^. To examine whether our decoders could detect the emotion-related abnormality among SUD individuals, we applied NERS-A and NERS-R to classify the distancing (one of reappraisal) strategies with negative emotions induced by negative pictures and also applied NNES to predict negative vs. neutral emotions. The data were provided by a previous study^68^ and newly collected data using a similar ER task paradigm and the identical scanner and sequence settings (see supplementary for details). Briefly, 48 health control participants (HC, 18 from our previous study^68^ and 30 newly collected) and 49 cannabis users (CU, 23 from our previous study and 26 newly collected) were included in the final analysis.

## Data availability

fMRI data used to train the signatures are available via GitHub at https://github.com/h-psy/fMRI_studies/tree/main/NERS. Other authors provided the fMRI data in the generalization cohort 1 via NeuroVault (https://neurovault.org/collections/16266)^28^, and fMRI data in the generalization cohort 2 via the OpenfMRI (https://openfmri.org/dataset/ds000140)^63^.

## Code availability

Code for analyzing data and generating figures is available via GitHub at https://github.com/canlab and https://github.com/h-psy/fMRI_studies/tree/main/NERS.

---

**Extended Data Table 1.** Classification performance between decoders on the validation cohort.

| Condition | Decoder | ACC (%) |  | Sens (%) |  | Spec (%) |  | Effect | *P* |
| --- | --- | --- | --- | --- | --- | --- | --- | --- | --- |
|  |  |  | SD |  | CI |  | CI | size |  |
| **Validation cohort (n=33)** |  |  |  |  |  |  |  |  |  |
| NV vs. NeutV | NNES | 100 | 0 | 100 | 100–100 | 100 | 100–100 | 3.55 | **2.33×10<sup>-10</sup>** |
|  | PINES | 91 | 5 | 91 | 80–100 | 91 | 79–100 | 1.16 | **1.40×10<sup>-6</sup>** |
|  | NERS-A | 70 | 8 | 70 | 53–85 | 70 | 53–85 | 0.95 | **0.035** |
|  | NERS-R | 3 | 3 | 3 | 0–10 | 3 | 0–10 | -33.03 | **7.92×10<sup>-9</sup>** |
| NA vs. NV | NNES | 48 | 8.7 | 48 | 31–66 | 48 | 32–67 | -0.3 | 1 |
|  | PINES | 36 | 8.4 | 36 | 20–54 | 36 | 21–53 | -0.12 | 0.1628 |
|  | NERS-A | 70 | 8 | 70 | 53–86 | 70 | 53–85 | 0.47 | **0.0351** |
|  | NERS-R | 61 | 8.5 | 36 | 19–53 | 36 | 21–53 | 0.61 | 0.2962 |
| NR vs. NV | NNES | 15 | 6.2 | 15 | 3–29 | 15 | 3–28 | -0.95 | **6.60×10<sup>-5</sup>** |
|  | PINES | 45 | 8.7 | 45 | 30–63 | 45 | 29–63 | -0.02 | 0.7283 |
|  | NERS-A | 70 | 8 | 70 | 54–85 | 70 | 53–85 | 0.72 | **0.0351** |
|  | NERS-R | 88 | 5.7 | 88 | 75–97 | 88 | 76–97 | 1.42 | **1.10×10<sup>-5</sup>** |

**Extended Data Fig. 1.**
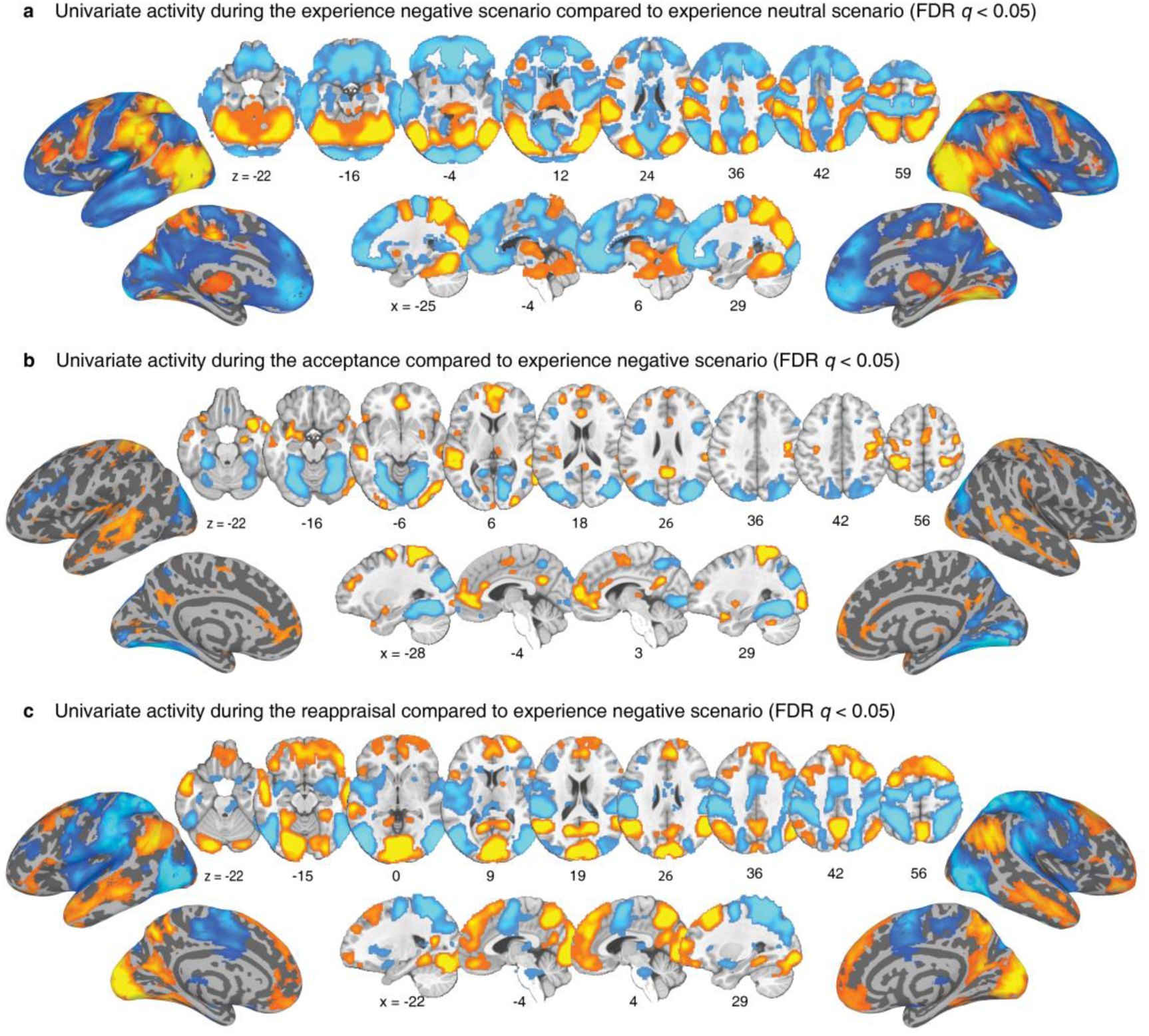
Brain activation from mass-univariate analysis. Contrast images representing the response to naturalistic negative scenario (view negative scenario (NV) vs. view neutral scenario (NeutV) (**a**), accept negative scenario (NA) vs. NV (**b**), and reappraisal negative scenario (NR) vs. NV (**b**) (FDR q<0.05).

**Extended Data Fig. 2.**
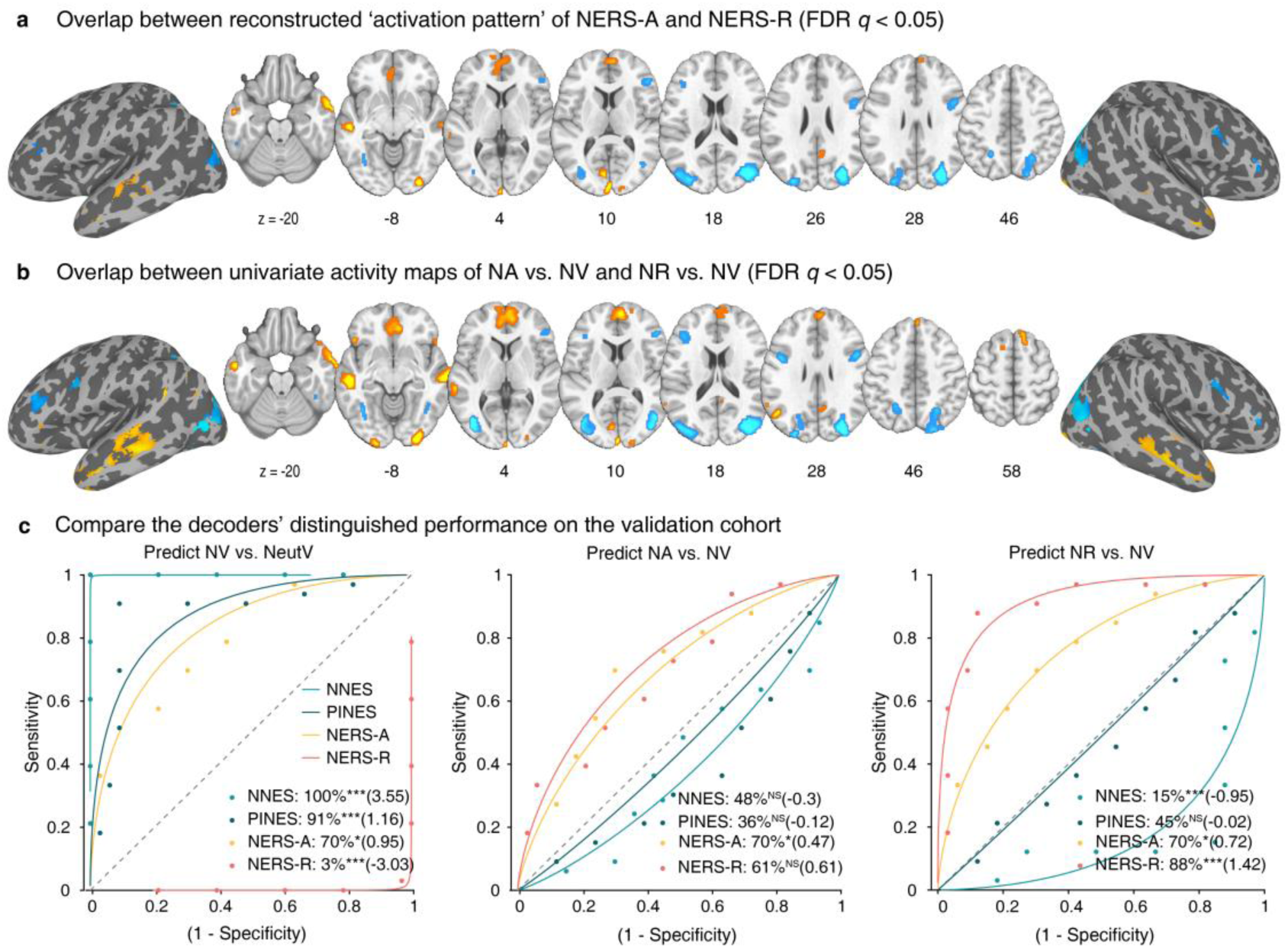
Common brain regions identified by conjunction analysis and comparison of decoders’ predictive performance. The conjunction of the FDR corrected (q<0.05) reconstructed ‘activation patterns’ between NERS-A and NERS-R (**a**) as well as univariate activity maps between NA vs. NV and NR vs. NV (**b**). **c**, Forced-choice classification accuracy and Cohen’s d indicate the NNES, NERS-A, and NERS-R could accurately predict the targeted mental processing during discrimination at NV vs. NeutV, NA vs. NV (left), and NR vs. NV (middle), respectively, based on the validation cohort. PINES could distinguish NV vs. NeutV but not ER strategies vs. NV.

**Extended Data Fig. 3.**
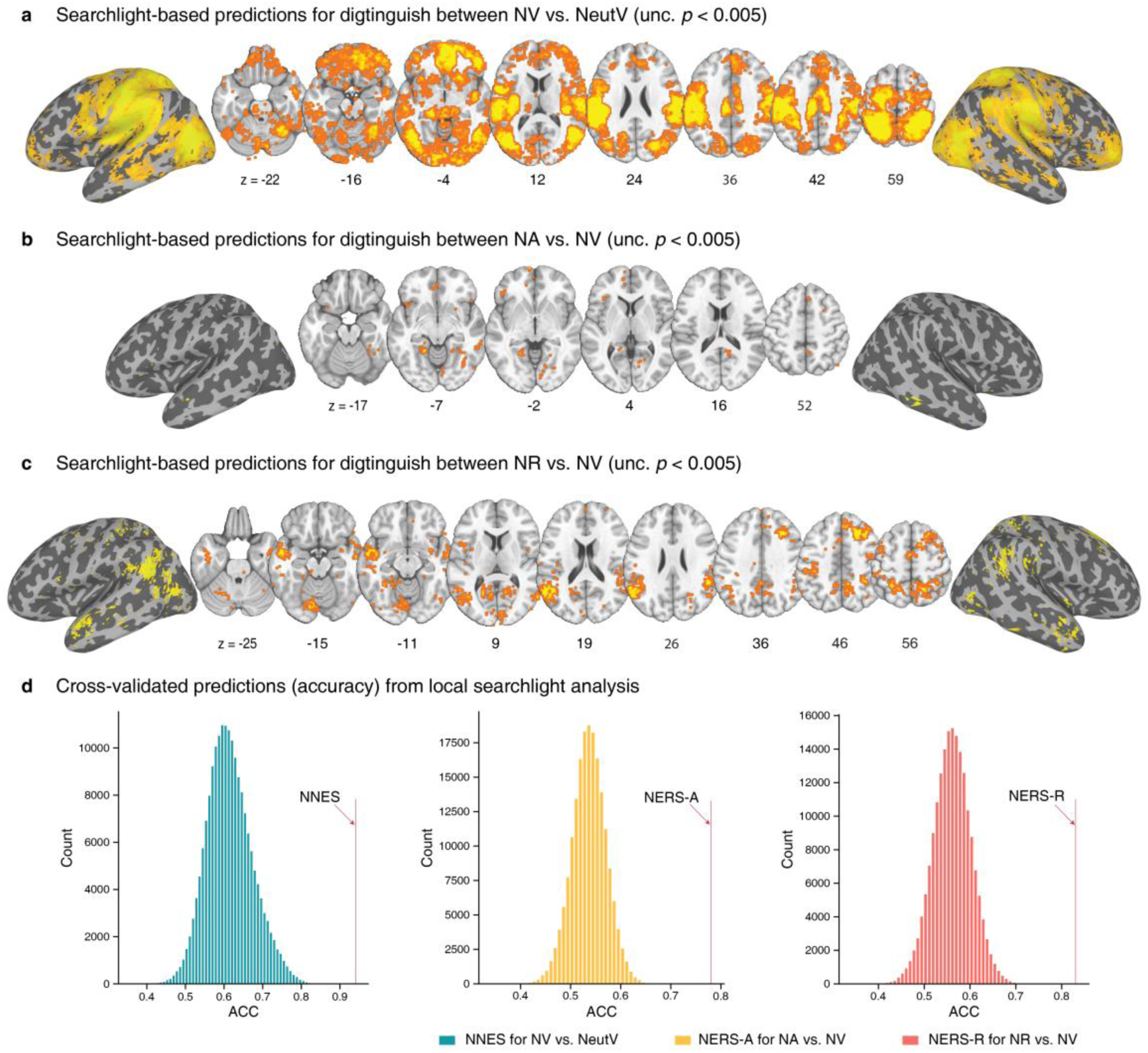
Searchlight-based predictions in the discovery cohort. Brain regions that significantly predict the response (**a**), acceptance (**b**), and reappraisal (**c**) to naturalistic stimuli that are revealed by searchlight-based analyses. **d**, Histograms: cross-validated predictions from local searchlights.

## Acknowledgments

This work was funded by the STI 2030 – major projects (Grant No. 2022ZD0208500, to DY, BB), National Natural Science Foundation of China (Grant No. 82271583, to BB), start-up/seed grant from the University of Hong Kong to BB, and was supported by the Sichuan Science and Technology Program (2023YFS0023, to BZ; 24NSFSC6564, to LL). Funders were not involved in the research design, the collection and analysis of data, the decision to publish, or the writing of the manuscript. We thank the authors of Bo, K. et al. and Woo, C.-W. et al. for providing data for the generalization cohorts in the present study, and we thank the authors of Chang, L. J. et al. for providing the PINES signature.

## Author contributions

HJ and BB were responsible for the research design and drafting of the manuscript. HJ, JH, KZ, BZ, LL, SF, LW, and XZ collected the data. HJ, ZF, and BB were responsible for the analysis and interpretation of data. XG, KMK, WZ, DY, and TY provided important suggestions and tools for the analyses and critically commented on the manuscript. B.B. supervised the project and acquired the funding. All authors meet the ICMJE’s four criteria for authorship and are responsible for revising the paper, approving the final version for publication, and ensuring the accuracy and completeness of the work.

## Notes

### Competing Interest Statement

The authors have declared no competing interest.

### Funding Statement

This work was funded by the STI 2030, a major projects (Grant No. 2022ZD0208500, to DY, BB), National Natural Science Foundation of China (Grant No. 82271583, to BB), start-up/seed grant from the University of Hong Kong to BB, and was supported by the Sichuan Science and Technology Program (2023YFS0023, to BZ; 24NSFSC6564, to LL). Funders were not involved in the research design, the collection and analysis of data, the decision to publish, or the writing of the manuscript.

### Author Declarations

This pre-registered study (discovery cohort: https://osf.io/s3bp2, validation cohort: https://osf.io/68jkw) was approved by the ethics committee of the University of Electronic Science and Technology of China and in accordance with the Declaration of Helsinki.

