## Supplementary for "Common and distinct neurofunctional signatures of emotion regulation strategies and their clinical translation in dynamic naturalistic contexts"

### Subjective negativity rating in the discovery and validation cohort

In the discovery cohort (n=59, utilized to develop the decoders) a one factorial repeated-measures analysis of variance (ANOVA) with the factor condition (NV, NeutV) revealed a significant main effect (*F* [1, 58]=1039.434, *P*=9.96×10^-39^, $\eta_{p}^{2}$=0.947; Fig.2a). Further simple effects analysis revealed that the negative view (NV) condition (6.52±1.21, Mean±SD) induced significantly higher negative emotional experience than the neutral view (NeutV) condition (1.58±0.55, *P*=9.96×10^-39^, 95% confidence interval (CI) [4.63, 5.25]). Given that the assumption of sphericity for one factor of the repeated-measures ANOVA of strategies (NV, negative–acceptance (NA), negative–reappraisal (NR)) was violated, the Greenhouse-Geisser correction was used, revealing a significant main effect (*F* [2, 95]=81.668, *P*=3.18×10^-19^, $\eta_{p}^{2}$=0.585). Further simple effects analysis revealed that both reappraisal (4.54±1.58) and acceptance (5.44±1.53) lead to significantly reduced subjective negative experience (NV: 6.52±1.21; NR vs. NV, *P*=1.54×10^-7^, 95% CI [-2.40, -1.55]; NA vs. NV, *P*=4.40×10^-16^, 95% CI [-1.49, -0.65]), indicating that both strategies allowed the participants to successfully regulate their negative emotional state. Reappraisal results in lower negative feelings than acceptance (*P*=2.02×10^-10^, 95% CI [-1.18, -0.62]). The successful emotion regulation was also mirrored in the self-report success rate of emotion regulation (9 - very successful, 1 - not successful at all, averaged success rating: discovery cohort: 7.51±1.01, validation cohort: 7.53±0.98).

In the validation cohort (n=33), one factor (stimulus type: NV, UV) repeated-measures ANOVA revealed a significant main effect (*F* [1, 32]=255.626, *P*=8.15×10^-17^, $\eta_{p}^{2}$ $\eta_{p}^{2}$=0.947). Further simple effects analysis revealed that the negative clips induced significantly stronger negative emotions than the neutral clips (NV vs. NeutV, 6.74±1.44 vs. 1.86±1.02, *P*=8.15×10^-17^, 95% CI [4.26, 5.50]). The assumption of sphericity from one factor repeated-measures ANOVA of strategies (NV, NA, NR) was violated, so the Greenhouse-Geisser correction was used, revealing a significant main effect (*F* [2, 52]=56.637, *P*=1.32×10^-12^, $\eta_{p}^{2}$=0.639). Both reappraisal and acceptance lead to significantly reduced subjective negative emotion (NR vs. NV, (4.43±1.83) vs. (6.74±1.44), *P*=7.07×10^-7^, 95% CI [-2.95, -1.66]; NA vs. NV, (5.3±1.53) vs. (6.74±1.44), *P*=1×10^-6^, 95% CI [-2.02, -0.86]). Reappraisal results in greater reduction than acceptance (*P*=2.4×10^-5^, 95% CI [-1.27, -0.45]).

### Table 1. Spatial similarity between model encoding and ROIs, as well as functional networks

| Name | Cosine Similarity | | |
| --- | --- | --- | --- |
|  | ACPT | REAPP | REACT |
| Prefrontal ROIs |  |  |  |
| dlPFC | 0.097 | 0.2216 | 0.0535 |
| vlPFC | 0.0761 | 0.0874 | 0.0539 |
| dmPFC | 0.1542 | 0.2251 | 0.009 |
| vmPFC | 0.0895 | 0.1494 | 0.0034 |
| OFC | 0.0471 | 0.1781 | 0 |
| ACC | 0.1034 | 0.0852 | 0.0244 |
| Functional networks |  |  |  |
| Visual | 0.4929 | 0.5694 | 0.4974 |
| Somatomotor | 0.4863 | 0.0419 | 0.1964 |
| dAttention | 0.3239 | 0.1412 | 0.5498 |
| vAttention | 0.2288 | 0.0572 | 0.2863 |
| Limbic | 0.1812 | 0.1531 | 0.0294 |
| Frontoparietal | 0.1267 | 0.2713 | 0.1104 |
| Default | 0.4282 | 0.5837 | 0.0807 |

dlPFC, dorsolateral prefrontal cortex; vlPFC, ventrolateral prefrontal cortex; dmPFC, dorsomedial prefrontal cortex; vmPFC, ventromedial prefrontal cortex; OFC, orbitofrontal cortex; ACC, anterior cingulate cortex; vAttention, ventral attention; dAttention, dorsal attention.

### Table 2. Prediction-outcome accuracy (mean ± std) of distinguishing between NA vs. NV with various numbers of voxels (for the Discovery Cohort)

| **Number**  **of voxels** | **Vis** | **SM** | **dA** | **vA** | **Limb** | **FP** | **DMN** | **PF** | **WB** |
| --- | --- | --- | --- | --- | --- | --- | --- | --- | --- |
| **50** | 58.83 | 56.88* | 59.13 | 55.06* | 55.2* | 57.99* | 58.47 | 58.24* | 58.73 |
|  | 3.56 | 3.52 | 3.58 | 3.38 | 3.56 | 3.68 | 3.68 | 3.67 | 3.93 |
| **150** | 61.95 | 58.81* | 61.86 | 54.99* | 54.93* | 58.57* | 60.37* | 59.32* | 62.18 |
|  | 3.19 | 3.31 | 3.21 | 2.88 | 3.16 | 3.04 | 3.27 | 3.29* | 3.41 |
| **250** | 62.84* | 59.12* | 62.55* | 55.24* | 55.42* | 59.18* | 61.51* | 59.82* | 65.17 |
|  | 2.96 | 2.99 | 2.75 | 2.76 | 2.83 | 2.79 | 2.99 | 2.86 | 3.4 |
| **500** | 62.71* | 59.41* | 63.74* | 55.95* | 56.18* | 60.15* | 62.7* | 60.58* | 68.91 |
|  | 2.62 | 2.61 | 2.44 | 2.53 | 2.35 | 2.57 | 2.85 | 2.71 | 3.18 |
| **750** | 62.15* | 60.15* | 64.48* | 56.36* | 56.44* | 60.42* | 63.27* | 61.01* | 70.91 |
|  | 2.36 | 2.36 | 2.4 | 2.51 | 2.17 | 2.34 | 2.75 | 2.49 | 3.06 |
| **1000** | 61.78* | 60.69* | 64.72* | 56.65* | 56.63* | 60.46* | 63.55* | 61.26* | 72.27 |
|  | 2.27 | 2.17 | 2.22 | 2.27 | 2.09 | 2.23 | 2.55 | 2.31 | 2.93 |
| **2000** | 61.09* | 61.68* | 65.32* | 56.95* | 56.86* | 60.71* | 64.04* | 61.8* | 74.6 |
|  | 1.86 | 2.02 | 1.88 | 2.08 | 1.7 | 1.87 | 2.23 | 2.01 | 2.5 |
| **4000** | 60.57* | 62.35* | 65.71* | 56.94* | 56.79* | 61.14* | 64.63* | 62.28* | 76.03 |
|  | 1.68 | 1.78 | 1.62 | 1.73 | 1.35 | 1.36 | 2.02 | 1.66 | 2.15 |
| **6000** | 60.38* | 62.65* | 65.99* | 57.07* | 56.66* | 61.21* | 64.94* | 62.59* | 76.43 |
|  | 1.54 | 1.65 | 1.37 | 1.46 | 0.99 | 1.15 | 1.9 | 1.49 | 1.99 |
| **8000** | 60.31* | 62.89* | 66.05* | 57.28* | 56.42* | 61.3* | 65.1* | 62.85* | 76.54 |
|  | 1.47 | 1.47 | 1.18 | 1.31 | 0.68 | 0.97 | 1.82 | 1.31 | 1.68 |
| **10000** | 60.23* | 63.01* | 66.07* | 57.47* | NA | 61.34* | 65.2* | 62.99* | 76.81 |
|  | 1.32 | 1.39 | 1.03 | 1.17 | NA | 0.83 | 1.73 | 1.17 | 1.58 |
| **14000** | 60.05* | 63.39* | 65.93* | NA | NA | 61.3* | 65.39* | 63.22* | 76.94 |
|  | 1.07 | 1.18 | 0.62 | NA | NA | 0.64 | 1.5 | 0.95 | 1.46 |
| **18000** | 59.72* | 63.67* | NA | NA | NA | 61.5* | 65.55* | 63.51* | 77.02 |
|  | 0.71 | 0.78 | NA | NA | NA | 0.47 | 1.37 | 0.75 | 1.46 |
| **25000** | NA | NA | NA | NA | NA | NA | 65.73* | 63.94* | 77.12 |
|  | NA | NA | NA | NA | NA | NA | 0.99 | 0.55 | 1.3 |
| **Full** | 59.32 | 63.56 | 65.25 | 57.63 | 56.78 | 61.86 | 65.25 | 64.41 | 77.97 |

Vis, visual network; SM, somatomotor network; dA, dorsal attention network; vA, ventral attention network; Limb, limbic network; FP, frontoparietal network; DMN, default mode network; PF, prefrontal cortex; NA, not applicable. * and # indicate that the prediction is significantly lower or higher as compared with the whole-brain model using the same number of voxels, separately (two-tailed t-test; P<0.05, Bonferroni corrected).

### Table 3. Prediction-outcome accuracy (mean ± std) of distinguishing between NR vs. NV with various numbers of voxels (for the Discovery Cohort)

| **Number**  **of voxels** | **Vis** | **SM** | **dA** | **vA** | **Limb** | **FP** | **DMN** | **PF** | **WB** |
| --- | --- | --- | --- | --- | --- | --- | --- | --- | --- |
| **50** | 64.64 | 61.1* | 65.25^#^ | 62.26* | 57.63* | 61.67* | 66.07^#^ | 61.09* | 64.42 |
|  | 4.41 | 3.65 | 4.17 | 4.1 | 3.91 | 4.22 | 4.41 | 4.04 | 5.05 |
| **150** | 69.5^#^ | 60.54* | 68.02* | 64.06* | 57.89* | 63.41* | 67.38* | 62.23* | 68.82 |
|  | 3.55 | 3.17 | 3.4 | 3.46 | 3.33 | 3.36 | 3.46 | 3.4 | 4.32 |
| **250** | 71.02* | 60.67* | 70.34* | 65.76* | 57.67* | 64.65* | 69.06* | 63.01* | 72.02 |
|  | 3.22* | 2.91 | 3.15 | 3.44 | 2.95 | 3.18 | 3.1 | 3.07 | 3.83 |
| **500** | 71.47* | 61.07* | 72.84* | 68.36* | 57.85* | 66.46* | 70.8* | 64.27* | 74.88 |
|  | 2.63 | 2.68 | 2.73 | 3.13 | 2.61 | 2.84 | 2.75 | 2.74 | 3.46 |
| **750** | 71.58* | 61.54* | 73.7* | 69.49* | 58.08* | 67.11* | 71.45* | 64.58* | 76.53 |
|  | 2.44 | 2.48 | 2.43 | 2.85 | 2.47 | 2.54 | 2.52 | 2.45 | 2.9 |
| **1000** | 71.52* | 61.73* | 74.14* | 70.22* | 58.21* | 67.43* | 71.95* | 64.62* | 77.22 |
|  | 2.23 | 2.33 | 2.22 | 2.69 | 2.39 | 2.44 | 2.4 | 2.24 | 2.69 |
| **2000** | 71.2* | 62.2* | 75.04* | 71.56* | 58.45* | 67.86* | 72.82* | 64.71* | 78.68 |
|  | 1.89 | 1.91 | 1.74 | 2.13 | 1.98 | 1.93 | 2.07 | 1.98 | 2.09 |
| **4000** | 70.84* | 62.66* | 75.45* | 72.61* | 58.46* | 68.02* | 73.35* | 65.06* | 79.59 |
|  | 1.58 | 1.49 | 1.34 | 1.65 | 1.64 | 1.53 | 1.7 | 1.72 | 1.88 |
| **6000** | 70.65* | 62.73* | 75.68* | 73.19* | 58.3* | 68.03* | 73.57* | 65.23* | 79.92 |
|  | 1.39 | 1.31 | 1.05 | 1.33 | 1.36 | 1.39 | 1.55 | 1.5 | 1.67 |
| **8000** | 70.59* | 62.81* | 75.76* | 73.62* | 57.99* | 68.07* | 73.68* | 65.38* | 80.14 |
|  | 1.24 | 1.18 | 0.93 | 1.16 | 1 | 1.21 | 1.42 | 1.39 | 1.56 |
| **10000** | 70.51* | 62.9* | 75.91* | 73.94* | NA | 68.2* | 73.71* | 65.6* | 80.35 |
|  | 1.19 | 1 | 0.76 | 0.94 | NA | 1.12 | 1.32 | 1.26 | 1.58 |
| **14000** | 70.47* | 62.93* | 76.14* | NA | NA | 68.45* | 73.78* | 65.8* | 80.73 |
|  | 1.01 | 0.8 | 0.35 | NA | NA | 0.85 | 1.15 | 1.09 | 1.44 |
| **18000** | 70.46* | 62.93* | NA | NA | NA | 68.85* | 73.76* | 65.88* | 80.94 |
|  | 0.83 | 0.55 | NA | NA | NA | 0.63 | 1.02 | 0.94 | 1.36 |
| **25000** | NA | NA | NA | NA | NA | NA | 73.55* | 65.98* | 81.28 |
|  | NA | NA | NA | NA | NA | NA | 0.7 | 0.68 | 1.23 |
| **Full** | 70.34 | 62.71 | 76.27 | 74.58 | 58.47 | 69.49 | 72.88 | 66.1 | 83.05 |

Vis, visual network; SM, somatomotor network; dA, dorsal attention network; vA, ventral attention network; Limb, limbic network; FP, frontoparietal network; DMN, default mode network; PF, prefrontal cortex; NA, not applicable. * and # indicate that the prediction is significantly lower or higher as compared with the whole-brain model using the same number of voxels, separately (two-tailed t-test; P<0.05, Bonferroni corrected).

### Table 4. Prediction-outcome accuracy (mean ± std) of distinguishing between NV vs. NeutV with various numbers of voxels (for the Discovery Cohort)

| **Number**  **of voxels** | **Vis** | **SM** | **dA** | **vA** | **Limb** | **FP** | **DMN** | **PF** | **WB** |
| --- | --- | --- | --- | --- | --- | --- | --- | --- | --- |
| **50** | 73.54* | 81.37^#^ | 79.5^#^ | 80.07^#^ | 62.15* | 74.11* | 70.12* | 72.99* | 77.96 |
|  | 4.47 | 4.56 | 4.24 | 4.49 | 3.97 | 4.41 | 4.16 | 4.13 | 4.95 |
| **150** | 80.56* | 86.1^#^ | 84.39 | 85.18^#^ | 63.27* | 77.57* | 71.65* | 74.89* | 84.47 |
|  | 3.74 | 3.38 | 3.23 | 3.49 | 3.46 | 3.47 | 3.32 | 3.27 | 3.87 |
| **250** | 82.61* | 87.84^#^ | 86.67 | 87.3^#^ | 63.95* | 79.31* | 73.36* | 75.99* | 86.91 |
|  | 3.12 | 2.82 | 2.61 | 2.85 | 3 | 3.11 | 3.04 | 3.04 | 3.16 |
| **500** | 84.14* | 89.19* | 88.41* | 89.2* | 64.44* | 81.34* | 75.27* | 77.35* | 89.74 |
|  | 2.45 | 2.21 | 2.03 | 2.3 | 2.51 | 2.7 | 2.57 | 2.6 | 2.97 |
| **750** | 84.65* | 89.7* | 88.92* | 89.97* | 64.58* | 82.06* | 76.09 | 78.04* | 90.96 |
|  | 2.25 | 1.95 | 1.82 | 2.09 | 2.23 | 2.34 | 2.2 | 2.26 | 2.35 |
| **1000** | 84.8* | 89.89* | 89.02* | 90.32* | 64.63* | 82.51* | 76.36 | 78.31* | 91.47 |
|  | 2.1 | 1.82 | 1.71 | 1.99 | 2.06 | 2.17 | 2.02 | 2.2 | 2.14 |
| **2000** | 84.86* | 90.65* | 89.37* | 90.98* | 64.58* | 83.26* | 76.79* | 78.65* | 92.77 |
|  | 1.87 | 1.52 | 1.41 | 1.63 | 1.65 | 1.53 | 1.51 | 1.85 | 1.63 |
| **4000** | 84.6* | 91.2* | 89.45* | 91.14* | 64.42* | 83.65* | 76.91* | 78.88* | 93.35 |
|  | 1.63 | 1.2 | 1.15 | 1.34 | 1.37 | 1.16 | 1.12 | 1.58 | 1.37 |
| **6000** | 84.41* | 91.51* | 89.6* | 91.22* | 64.21* | 83.79* | 76.84* | 78.96* | 93.69 |
|  | 1.44 | 1.13 | 1.01 | 1.12 | 1 | 0.96 | 1 | 1.48 | 1.24 |
| **8000** | 84.24* | 91.81* | 89.66* | 91.13* | 63.86* | 83.84* | 76.75* | 79.02* | 93.93 |
|  | 1.3 | 1.04 | 0.91 | 0.97 | 0.48 | 0.74 | 0.87 | 1.33 | 1.11 |
| **10000** | 84.12* | 92.03* | 89.75* | 91.01* | NA | 83.89* | 76.66* | 79.07* | 93.89 |
|  | 1.14 | 0.92 | 0.79 | 0.84 | NA | 0.59 | 0.82 | 1.24 | 1.08 |
| **14000** | 83.96* | 92.33* | 89.93* | NA | NA | 83.9* | 76.53* | 79.07* | 94.01 |
|  | 0.9 | 0.75 | 0.41 | NA | NA | 0.27 | 0.79 | 1.12 | 1.08 |
| **18000** | 83.74* | 92.66* | NA | NA | NA | 83.9* | 76.37* | 79.07* | 94.17 |
|  | 0.71 | 0.62 | NA | NA | NA | 0 | 0.73 | 0.99 | 1.09 |
| **25000** | NA | NA | NA | NA | NA | NA | 76.03* | 78.86* | 94.11 |
|  | NA | NA | NA | NA | NA | NA | 0.57 | 0.72 | 0.94 |
| **Full** | 83.05 | 93.22 | 89.83 | 90.68 | 63.56 | 83.9 | 75.42 | 78.81 | 94.07 |

Vis, visual network; SM, somatomotor network; dA, dorsal attention network; vA, ventral attention network; Limb, limbic network; FP, frontoparietal network; DMN, default mode network; PF, prefrontal cortex; NA, not applicable. * and # indicate that the prediction is significantly lower or higher as compared with the whole-brain model using the same number of voxels, separately (two-tailed t-test; P<0.05, Bonferroni corrected).

### MRI data acquisition and preprocessing of the clinical application cohort

**Participants**

Data from eighteen male healthy control (HC) participants and twenty-three male heavy recreational cannabis users (CU) were obtained from our previous study^1^. For the present study, we increased the sample size and recruited thirty-six healthy male HC participants and thirty-six male CU participants according to comparable criteria to our previous study^1^.

For all recruited participants, exclusion criteria included (1) a history of psychiatric disorder (measured by the Mini-International Neuropsychiatric Interview, M.I.N.I.^2^), except for cannabis use disorder, (2) regular/current use of psychoactive/cardiovascular medication, (3) positive urine screen for the substances cocaine, methamphetamine, amphetamine, or methadone, or (4) breath alcohol level >0.00. For CU, additional inclusion criteria included (1) long-term regular cannabis use (use on over 200 occasions, in the new sample cannabis users had used cannabis regularly for 77.90±51.53 months; during the past year on 25.35±6.37 days per month) additional exclusion criteria included (1) reported having used other illicit substances on > 50-lifetime occasions (2) use of cannabis or other substances in the 24 hours before the experiment. For HC, additional exclusion criteria included a positive of tetrahydrocannabinol (THC) testing on the day of the fMRI scanning.

Three HC and seven CU newly collected participants were excluded due to excessive head motion (> 3 mm or 3°); three HC and three CU were excluded due to being detected as outliers based on their emotion regulation success (defined as the mean decrease of the negative affect rating for reappraisal (“distance”) trials relative to ratings for emotional reactivity (“spontaneous_negative”)^1^. Thirty HC (26.1 ± 4.8 years old) and twenty-six CU newly recruited participants (26.6 ± 5.8 years old) were included in the final analysis, leading to a total of 48 HC and 49 CU participants were included as the clinical application cohort. No significant group difference in years of age and education. All participants provided written informed consent.

**Stimuli and Paradigm**

The stimuli and paradigm are similar to our previous study^1^, except the current study uses a negative affect rating scale from 0 to 100, in which 0 represents no negative, 50 represents a moderate negative, and 100 represents a strong negative.

**MRI data acquisition and preprocessing**

MRI data were acquired using an identical scanner and sequence settings to our previous study^1^. Use the same preprocessing script as we did on the discovery and validation cohort (only modifying the parameters based on the sequence setting).

2.	Sheehan, D. V. The Mini-International Neuropsychiatric Interview (M.I.N.I.): The Development and Validation of a Structured Diagnostic Psychiatric Interview for DSM-IV and ICD-10.
